# Graphical Models of Pandemic

**DOI:** 10.1101/2021.02.24.21252390

**Authors:** Michael Chertkov, Ruby Abrams, Amir Mohammad Esmaieeli Sikaroudi, Mikhail Krechetov, CNP Slagle, Alon Efrat, Radoslav Fulek, Eyal Oren

## Abstract

Both COVID-19 and novel pandemics challenge those of us within the modeling community, specifically in establishing suitable relations between lifecycles, scales, and existing methods. Herein we demonstrate transitions between models in space/time, individual-to-community, county-to-city, along with models for the trace beginning with exposure, then to symptom manifest, then to community transmission. To that end, we leverage publicly available data to compose a chain of Graphical Models (GMs) for predicting infection rates across communities, space, and time. We’ll anchor our GMs against the more expensive yet state-of-the-art Agent-Based Models (ABMs). Insight obtained from designing novel GMs calibrated to ABMs furnishes reduced, yet reliable surrogates for the end-to-end public health challenge of community contact tracing and transmission. Further, this novel research transcends and synergizes information integration and informatics, leading to an advance in the science of GMs. Cognizance into the data lifecycle using properly coarse-grained modeling will broaden the toolkit available to public health specialists, and hopefully empower governments and health agencies, here and abroad, in addressing the profound challenges in disease and vaccination campaigns confronting us by COVID and future pandemics.

In this proof of principle study, focusing on the GM methodology development, we show, first, how static GM of the Ising model type (characterized by pair-wise interaction between nodes related to traffic and communications between nodes representing communities, or census tracts within a given city, and with local infection bias) emerge from a dynamic GM of the Independent Cascade type, introduced and studied in Computer and Networks sciences mainly in the context of the spread of social influences. Second, we formulate the problem of inference in epidemiology as inference problems in the Ising model setting. Specifically, we pose the challenge of computing Conditional A-posteriori Level of Infection (CALI), which provides a quantitative answer to the questions: What is the probability that a given node in the GM (given census tract within the city) becomes infected in the result of injection of the infection at another node, e.g. due to arrival of a super-spreader agent or occurence of the super-spreader event in the area. To answer the question exactly is not feasible for any realistic size (larger than 30-50 nodes) model. We therefore adopt and develop approximate inference techniques, of the variational and variable elimination types, developed in the GM literature. To demonstrate utility of the methodology, which seems new for the public health application, we build a 123-node model of Seattle, as well as its 10-node and 20-node coarsegrained variants, and then conduct the proof of principles experimental studies. The experiments on the coarse-grained models have helped us to validate the approximate inference by juxtaposing it to the exact inference. The experiments also lead to discovery of interesting and most probably universal phenomena. In particular, we observe (a) a strong sensitivity of CALI to the location of the initial infection, and (b) strong alignment of the resulting infection probability (values of CALI) observed at different nodes in the regimes of moderate interaction between the nodes. We then speculate how these, and other observations drawn from the synthetic experiments, can be extended to a more realistic, data driven setting of actual operation importance. We conclude the manuscript with an extensive discussion of how the methodology should be developed further, both at the level of devising realistic GMs from observational data (and also enhancing it with microscopic ABM modeling and simulations) and also regarding utilization of the GM inference methodology for more complex problems of the pandemic mitigation and control.

---

COVID-19 has acutely demonstrated the difficulty in both predicting and neutralizing the spread of pandemics. Critical challenges remain both in the scarcity of coherent data lifecycle modeling and reconciliation between global tactics and strategies. Indeed, the ongoing crisis in America exhibits the frailties both in key infrastructure and mitigation strategies. To address said global challenge of connectivity, we must model conciliation within the scopes *of space, time*, and *scale*, examples including individual to community, county to the city, and from the moment an infectious molecule first enters our bodies, to days of disease development and to community transmission. This is one of the most puzzling scientific challenges of our age with dramatic consequences for the future, with implications transcending the current scope and domain. We believe the solution nestles within a synthesis of epidemiology, applied mathematics, statistics, data science, and artificial intelligence.

We hope to stimulate progress by crowd-sourcing our models to a broad community of researchers. We expect to populate into the existing Public Health (PH) landscape what we believe to benovel Artificial Intelligence (AI), Data Science (DS) and Machine Learning (ML)-related mathematical and application-inspired concepts, namely Graphical Models of Pandemics (GMPs); these mathematical objects in principle can express the probabilistic spatio-temporal spread of pandemics to any degree of accuracy desired, contingent upon the complexity of the model. By contrast, the GMPs we introduce herein ought be viewed as

- reduced macroscopic models, as opposed to Agent-Based Models (ABMs) or other higher fidelity models higher-fidelity models (as an example, GMPs may resolve to a neighborhood, whereas ABMs resolve to individuals; of course, GMPs could, in principle, resolve to individuals, though the model suffers higher computational complexity; expected trade-offs strike the heart of our work here)
- efficient in computing probabilistic predictions (for instance offering the marginal probability heat map for the city neighborhoods to transition from the current/prior state of infection to the projected/a-posteriori state in two weeks)
- falling into one of
  i. dynamic GMs of the cascading type [59, 74, 77, 100, 108], parameterized by pairwise probabilities of the infection transmission between neighborhoods within a supplied unit of time, and the graph of the neighborhoods and their pairwise connections; or
  ii. static GMs capable of querying, for instance, stabilized randomized heat maps given an exogenous injection (such as the massive super-spreader event following Thanksgiving) and the city’s a-priori state of infection.
- data-driven; that is, our GMPs extract from open sources such as pandemic data repositories, the U.S. Census and the Geographical Information System (GIS), and mobility data; GMPs
  i. combine this input data with preselected parameters and output data cross-validated with corresponding ABMs, and
  ii. corroborate results with estimations provided by the subject matter experts (SMEs) in Public Health

We believe advantages of GMPs delineated above uniquely position our design and team to play a role in the resolution of both this pandemic and future such events.

More specifically, COVID-19 challenges modelers and healthcare professionals with

- asymptomatic development of the earliest stages,
- fast and only partially known mechanisms of person-to-person spread (such as airborne vs. droplet vs. fomite),
- significant uncertainty in how the pandemic spreads between communities, enormous geographical variations, and
- strong dependence on social and cultural differences.

Certainly, our understanding of the data lifecycle of pandemics has evolved since March of 2020, leading to more coherent models after building upon the incipient

- long-lasting “hammer-and-dance” mitigation [106],
- first public health-informed spatially aggregated data [92], and
- high-resolution [51].

The tremendous suffering COVID-19 has caused during 2020 and early 2021[101], the immense vulnerability the disease has exposed within our medical and economic infrastructure, and the computational complexities described above furnish deep motivation to research and collaborate the full efficacy of GMPs. Following conventional data-driven modeling, We emphasize the data lifecycle herein. Siphoning multiple data-sources available due to rapid global spread and widespread mortality of COVID-19, we will develop models to simulate, somewhat specifically, said spread, snapshot inference and estimation, and, more generally, mitigation of pandemics heretofore unseen. Despite our theoretical focus here, we expect validation of the models to follow in the future from a growing compendium of relevant data resources, so far including

- historical data [7] repositories of global data on confirmed cases,
- deaths, such as [3, 5],
- COVID-19 templates, metrics, and evaluations [8, 13],
- lists of medical repositories such as CT scan [107],
- widgets to test scenarios [6], and
- information on many COVID-19 tracking apps and websites [80].

Figure. 1 illustrates a general schematic of the model components and respective computations, the specifics of which we’ll discuss below. We can characterize said hierarchy with a comprehensive investigation of the epidemiology data lifecycle, namely with efficient model reduction endemic to graphical models. In some additional details and with pointers to other related efforts:

→ Agent-Based Models (ABMs) constitute the backbone of quantitative epidemiological models. We review the state-of-the-art in the ABMs in section I B with a focus on the design and validation of the reduced models of pandemics.
→ Sections II and III constitute the core of the manuscript. Here we describe our approach for design and inference on a spatio-temporally reduced model, coined Graphical Models of Pandemics (GMPs), which are lighter than an ABM of comparable expressive power.

**FIG. 1:**
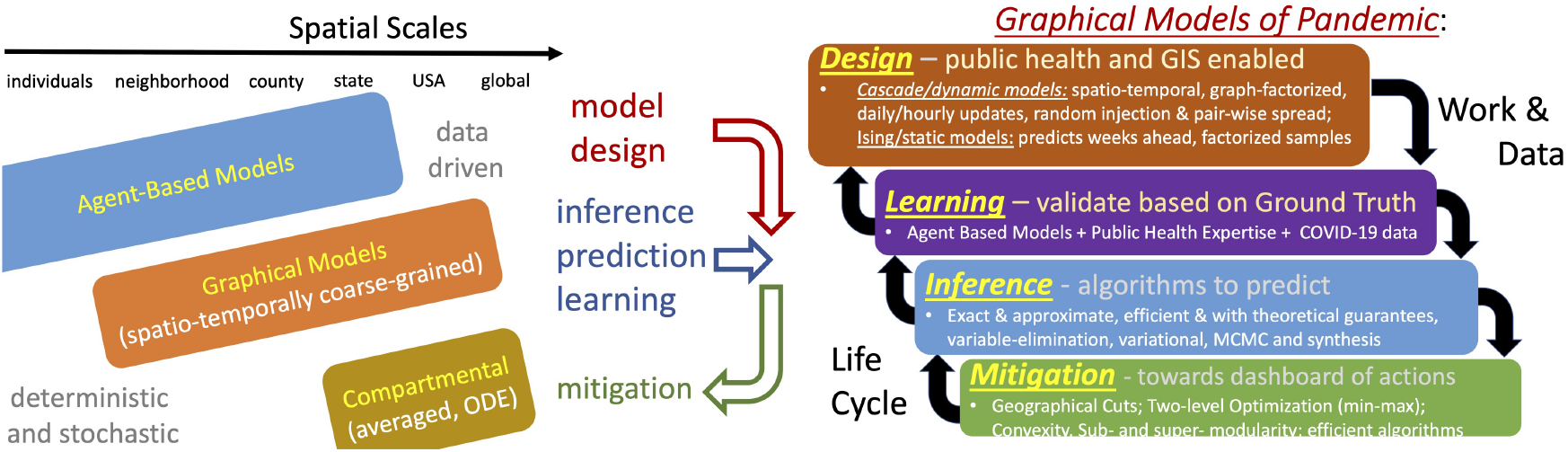
Left: Hierarchy of Scales, Models and Problems. Right: A condensed illustration of the logical connections and highlights of our inference approach, along with speculative inference, learning and control/mitigation challenges to be addressed in the follow up work.

We define more clearly the GMP in Section II, adapting data structures and algorithms from the computer science literature to model the spread of information, for instance rumors or news, between communities [59, 74, 77, 100, 108]. Such Dynamic Graphical Models (DGM), for instance the Independent Cascade Model (ICM) of [74], offer a predictive probabilistic approach for tracking samples of the system/network epidemiological status. Succinctly, we condition these models upon an initial state of the system infection, for instance an injection by a super spreader event, after which they output a terminal epidemic status reached within the period and geography of interest, say a few weeks after the initial injection of the infection in the system, along with a respective behavior spatially.

Section III exhibits an analysis of the posterior probability distribution of possible terminal states. Suppose we model a city with an undirected graph 𝒢 = (𝒱, ℰ), wherein each node *a* ∈ 𝒱 represents a census tract or neighborhood, and each edge *ab* ∈ ℰ corresponds to a daily route of persons traveling between the pair of census tracts corresponding to the endpoints of said edge. Following the Ising model from statistical physics, we assign to *a* ∈ 𝒱 the positive and negative unit to a respective pair of states; more technically, *x*_*a*_ = +1 indicates the state **R**emoved, meaning a significant percentage of the population experienced the infection over some prescribed observation period, and *x*_*a*_ = *−* 1 corresponding to the **S**usceptible state over the same period. Thus, we can represent the probability distribution of observing the city in the ***x*** state in the terminal state (when the spread, caused by the super-spreader incident, stops) with the following minimal Graphical Model of Pandemic (minGMP), or Ising Model of Pandemics:

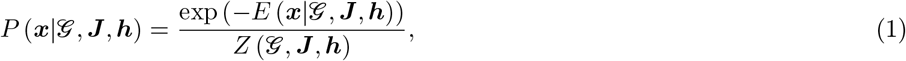

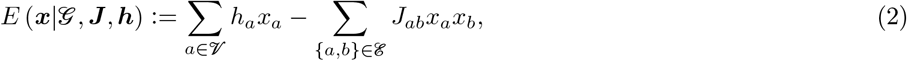

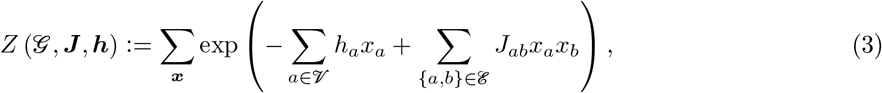

where

- *P* (***x***|𝒢, ***J***, ***h***) is the probability of observing the city in the ***x*** state by the end of the month,
- ***h*** and ***J*** are node-local bias and pair-wise interactions respectively,
- *E* (***x***|𝒢, ***J***, ***h***) is the energy of the state,
- *Z* (𝒢, ***J***, ***h***) is the partition function,
- ***h*** represents the initial pattern of infection, for example, due to a super-spreader event at a node or group of nodes, and also level of the node immunity, via population health, level of restrictions, and so on, and
- ***J*** represents the intensity of the pair-wise interaction between nodes due to the inter-community travel. Then we may evaluate Conditioned A Posteriori Level of Infection (CALI)

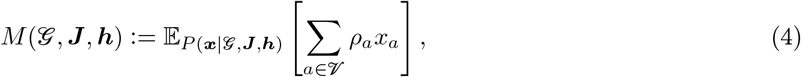

as an expectation over the probability distribution of ***x*** by the end of the period – red/green when it approaches +1*/−* 1 respectively) – conditioned upon the population density over the census tracts represented by ***ρ***, and the communities’ intro-status and interactions represented by ***h*** and ***J***, respectively.

Sections III and IV respectively propound our most important technical contributions herein and examples thereof. We delineate these results below:

- Section III A contains a derivation of the Ising model of pandemic (1) from its dynamic counterpart – we regularize the ICM to coerce graph-local properties on the terminal distribution.
- Section III B describes both the computations and exposition of significance for related objects, such as the system aggregate CALI index (22) and its graph non-homogeneous “heat-map” variation, coined A Posteriori Level of Infection (ALI).
- Section IV demonstrates how we combine the General Information System (GIS) data and synthetic but quasi-realistic epidemiological situation of interest to build an exemplary Ising model for the city of Seattle; further, the section suggests efficient inference algorithms of the variable elimination type [20, 21] in computing CALI index, the ALI heat-map, and other related objects of interest, such as higher order correlation functions, making more detailed probabilistic predictions on the spatiotemporal spread of pandemic.

Section V summarizes the results herein. Section VI offers path forward insights into learning GMPs from available data more concretely than how we define them in the manuscript, namely by incorporating and developing modern machine learning approaches, such as pseudo-loglikelihood, interaction screening, and artificial neural networks. We’ll also discuss here how to validate GMPs with the publicly available health input, COVID-19 data, and the ground truth data extracted from the high-resolution runs of relevant ABMs. Finally, we’ll suggest how inference and learning of the GMPs, both difficult computationally, nonetheless represent a keystone in mitigation strategies, approaches, and algorithms.

## I. INTRODUCTION: SETTING THE STAGE FOR THE GRAPHICAL MODELS OF PANDEMIC

### A. Uncertainties in Modeling Pandemics: Public Health Introduction

Multiple uncertainties render prediction of the spread of pandemics extremely difficult, an observation all the more concrete as we cope with COVID-19.

By now, researchers and laypersons alike recognize the significance of estimating the *R*_0_-parameter, a representation of the average number of people infected by one infectious individual. An *R*_0_ larger than unity suggests the infection spreads, whereas *R*_0_ less than unity inevitably leads to an end to an epidemic. The parameter *R*_0_ appears either explicitly or implicitly in all the models, hence its key position in the hierarchy of parameters relevant in modeling and mitigating pandemics. Moreover, the public health sciences recognized this importance generating one of the first predictions of *R*_0_ for COVID-19 in January of 2020 [90], leveraging then current case numbers, along with person-to-person spread rates superimposed from earlier studies of the Middle Eastern Respiratory Syndrome (MERS), another coronavirus infection. This and other early estimates exceeded two, and were more typically in the two-to-3.1 range. Indeed, accurate estimates of *R*_0_ can provide a necessary and first step in a global mitigation strategy, yet decent estimation poses a number of challenges, chiefly of which are mysterious biochemical characteristics of the virus, along with many other factors briefly discussed below. First, we cannot stress enough that the interpretability, popularity, and ensuing importance of *R*_0_ with respect to a given pandemic still constitutes an *averaged* parameter, capturing the difficulty in representing a population of geographies, points in space and time, and transmission modalities by a single mean value. This difficulty appears throughout the statistical and recent COVID-19 studies, raising some question of viability of *R*_0_ as a single measure of importance. It nonetheless provides a starting position, and accurate estimation remains a paramount challenge [42].

Articulating and quantifying uncertainties contributing to *R*_0_ and other characteristics of pandemic models will aid our approaches to quantifying these based on available data; we aim to predict the data lifecycle of a pandemic spread weeks ahead on the geographical scale of, say, a big city. Therefore, we’ll focus upon listing major uncertainties in modeling how the pandemic enters a model city and how it spreads within the city.

### Exogenous Injection

We know neither the point in time nor location in which the infection injects into the city. Possibilities include air-travelers, returning residents, and/or visitors coming by cars. To estimate the rate and locations of infection injection, we’ll consider the volume of arrivals and respective distributions over communities comprising final destinations.

### Internal Spread

Modeling transmission of the infection within the modeled city presents greater uncertainty than that of injection of the infection from the outside. Below we provide an incomplete list of relevant effects.

- Transmission occurs from person-to-person when people meet. The endemic challenge in predicting human behavior, particularly with daily travel routes through the city and levels of assumed protection, partially obscures propagation throughout the geographic unit.
- Touch-surfaces can transmit the disease, as we’ve discovered that the virus can survive in vitro for some number of hours; the count of such surfaces, exposure rates, time-scales associated with travel and visitation volume, and, again, erratic self-protection and retail/business enforcement rates further propagate uncertainty.
- Airborne transmission of COVID-19, especially indoors, varies highly. Estimations of the airborne disease transmission aimed at deriving an indoor safety guideline showed dependence of the “cumulative exposure time” on the rates of ventilation and air filtration, dimensions of the room, breathing rate, respiratory activity and face-mask use of its occupants, and infectiousness of the respiratory aerosols [30].
- As mentioned above, the strength of transmission fluctuates as it varies significantly from one individual to another. The modeling is complicated by the fact, recently discovered through phylogenetic analysis of the COVID-19 samples collected over many infected individuals [117], that the disease spread is extremely intermittent – a small percentage (10% or less) of individuals cause significant percentage (90% or more) of the secondary cases. Moreover, COVID-19, like most dangerous infections, may transmit asymptomatically, therefore rendering tracing extremely important but also quite challenging.
- Dependence of the transmission rate on temperature, UV and humidity, and other environmental factors, further expands the uncertainty. We remain ignorant as to whether seasonality will alter its spread and justify restarting economic activities. Ultraviolet light, in particular, seems correlated with decreased disease growth rate relative to other analyzed factors [95].
- We to account for spatio-temporal epidemiological inhomogeneity of the city, e.g. related to spatial diversity of *R*_0_, which certainly cannot be considered as the single parameter characterizing the status of the entire city [107].

Many other uncertainty factors originate from specifics of the national, regional, and individual responses to the pandemic.

The three **T**s in the non-pharmaceutical public health response – **Testing, Treating**, including preventing large gatherings, voluntary or enforced social distancing, curfews, road blocks, closure of businesses, and so on, and contact **Tracing** influence very significantly how we model the pandemic spreads; see [51], [92] and references therein for early illustrations of difficulties in modeling and uncertainty in the results of the COVID-19 spread. Recent work (https://www.thelancet.com/journals/laninf/article/PIIS1473-3099(20)30785-4/fulltext) illustrates the impact of both the introduction and lifting of non-pharmaceutical intervention policies on SARS-CoV-2 transmission

Representing the effects of vaccination on both external injection and internal spread over the model geography (for example of the model city) adds yet more uncertainty to an already very significant modeling challenge. While modeling prior pandemics [49], researchers attempted to account *post factum* for vaccination, a critical factor in controlling COVID-19 (through broader coverage as well as efficacy) as vaccines gradually become available.

Irrespective of these technical challenges, our species understandably hopes to survive this pandemic, and thus we as researchers bear an incumbency to answer and resolve as many of the relevant questions as possible; we intend to address the following in the mansucript on the scale of a modeled city are:

- Given a super spreader event (an exogenous injection) or number of these incidents, how do we predict whether the infection spreads? How fast, far, and intense?
- To monitor the spread, what types and how much data must we collect?
- By way of mitigation, how fast and when should we act, either individually, or at the level of county, city or state leadership? Can and should we construct a dashboard presentation of the model outcome for decision making?
- How can we best leverage what likely will be a dearth of spatio-temporal data to approximate the spread?
- Finally, and summarizing, how can we build a reliable and scalable **data lifecycle model of the pandemic**?

In summary,in the bigger picture of efforts to address the aforementioned challenges and questions, we aim by devising reduced models of pandemics based on the field of Graphical Models, a suite of modeling algorithms and data structures originating from Computer Science, Discrete Math, Artificial Intelligence, Statistical Physics, Data Science, and Machine Learning. We intend to complement the currently available Agent-Based type models that suffer from heavy implementation and computational complexity, with the global objective of offering a more efficient mitigation strategy for modeling pandemics [9].

### B. High-Resolution Models of Pandemic: Agent Based Models

The Agent Based Models (ABMs), introduced in epidemiology in 2004-2008 [49, 52, 53, 56, 63, 88], has complemented the compartmental models [24, 67, 75, 109]. Using ABMs, even though not exclusive to epidemiology [44, 120], became a breakthrough in the field, because they allowed for significant improvement in the quality of predictions, especially in the spatio-temporal resolution of how the disease spreads and how one can mitigate its spread. The models became and remain a core part of the epidemiology data life-cycle. (See for instance [89] for recent bibliography.) The ABM models provide a detailed prediction of how epidemics and pandemics spread within counties, cities, and regions. A majority of the country-, city- or county-scale testbeds testing various mitigation strategies are resolved nowadays with ABMs. In particular, recently ABMs have been used extensively to inform public health in (non-pharmaceutical) interventions against the spread of COVID-19 [4, 48, 51, 73, 94], among many other applications. Epidemiological ABMs have also gotten their share of criticism by public health experts [51, 112, 113], for instance for injecting too many, and often unrealistic assumptions. Still, they are considered to be most useful in informing decisions regarding mitigation and suppression measures in cases when ABMs are accurately calibrated [94].

The issue of calibration is central to what we propose. ABMs are clearly over-modeled. For example, the open-source ABM solver FLUTE [33] works with data that are acquired through Geographic Information Systems (GIS) on the scale of census tracts or communities, which is a very reasonable scale of spatial resolution to understand the dynamics of pandemics on a local scale. FLUTE populates each of the communities with thousands-to-millions of inhabitants in order to account for their daily patterns of travel. We believe that constructing effective Graphical Models of Pandemics with community-scale spatial resolution and then modeling pairwise (and possibly higher order) epidemic interactions between communities directly, without introducing the thousands-to-millions of dummy agents, will complement (as discussed in the next paragraph), but also improve upon ABMs by being more efficient, robust and easier to calibrate.

Dynamic and Static Graphical Models of Pandemics will be discussed in the following Sections. It is important to emphasize upfront that the models will need to be calibrated, or in other words need to be trained. The ultimate goal will be in training the Graphical Models on real-world data. However, we first aim to validate the GM methodology on ABMs, which are considered as synthetic models representing the ground truth.

We choose to work with FLUTE [33] because it meets the requirements outlined above and is also open access. However, our Graphical Models of Pandemic will not be linked specifically to FLUTE software. Moreover, we will be aiming in the future to adapt and verify our Graphical Models on other ABMs, particularly those developed and maintained by the MIDAS community [9].

In the remainder of this Section, and having outlined significance of the ABMs for this manuscript and broader efforts in mind, we briefly describe epidemiologically significant degrees of freedom embedded in the ABMs.

ABM constitutes a bottom-up modeling approach which forms a simulation by a collection of detailed small elements called agents. ABM is useful when the global behavior of a system is unknown but the interaction between agents is partially known and possibly complex, mimicking reality. Agents interact with environments and themselves to make decisions and traverse in the simulation environment.

The ABM models account for (a) geographical maps; (b) daily agent routines, (c) when households are resolved; (d) age, job, ethnicity of the agents. Typical input settings of the ABMs includes current status of the highly resolved system (millions of agents) and pre-defined rules of agent operations. ABMs outputs trajectories and change of status of the millions of agents over multiple days. This very detailed output may then be processed to extract aggregated characteristics of interest, like level of infection over the ABM geographical location over the time period of the simulation. It is clear that running a single ABM run is extremely demanding. Top federal HPC resources available in the USA (DOE, NSF, NIH) and world-wide are utilized heavily to run the high-resolution ABMs. Notice also that to test different scenarios (such as how initial infection lands in the city of interest) and mitigation strategies (pharmaceutical, if a vaccine is available, as well as non-pharmaceutical, like road or school closure, and so on.) will require running already extremely heavy computations thousand-to-millions number of times. Finally, it is important to stress that ABMs may also provide powerful simulation environments where different types of public health interventions, programs, and policies can be tested when more traditional public health outcome studies are not possible [81].

There are some subtle differences in the ABMs used in the community of researchers advancing the state of the art in ABM, in particular if the model works with an agent or actual individual. Three basic approaches have been tested: the UCL and U of Pittsburgh approach [51–53], also continued by many other [69, 73], model infectious interaction via spatial kernels; the University of Washington/LANL approach [56, 88], later resulted in FLUTE [33], consists in allowing individuals to move according to a number of prescribed trajectories; and Virginia Bionformatics Institute model [49] sets up trajectories based on samples of real people. ABMs were adopted to model different interaction scenarios between agents [31, 73], as well as different types of interactions of agents with the environment. The three approaches were juxtaposed to each other in [63] and they now form a backbone for many other versions and hybrids maintained and developed by the MIDAS community [9, 39].

Providing more information on our initial ground-truth model: FLUTE is an open-source library written in C/C++ that simulates individuals from city scale to country scale [33]. Geographical Information Systems (GIS) have the essential role to make the simulation possible. Individuals are grouped into households, neighborhoods, communities, and census tracts. Daily activities of people are simulated based on GIS data of travel patterns inside cities. The GIS data consists of employment information and demographic information of census tracts. Additionally, travel patterns among cities are considered. FLUTE can simulate different scenarios such as lock-down policies for a tract, closure of schools, individual quarantine, and so on. FLUTE was designed for influenza pandemic therefore allowing to simulate vaccinations and antiviral medications. The library utilizes Open MPI for parallel processing. FLUTE creates synthetic population hierarchy of “mixing” groups (household, neighbors, schools, etc); two time steps a day, infectious for 6 days (the case of influenza); travel (long-distance, short term); infection, contact probability (the same site), *R*_0_-related; simulated different types of interventions [mitigation – site-dependent rate of infection, isolation, etc]; entire US, 280 mln individuals, 60-90 days, also individual cities - Seattle (100+ census points), Los Angeles (2000+ census points). We use Seattle for illustration in the Figure above, showing in its left and right sub-figures respectively the travel pattern between census tracts of Seattle and spatial distribution of the population density, where the green disks represent population of census tract and the blue disks represent population densities of the census tract.

## II. CASCADE MODEL OF PANDEMIC: FROM DYNAMIC TO STATIC GRAPHICAL MODEL

We are interested in studying the spread of a virus in a network of people, communities, cities, and so on. To represent dynamics of the infection spread between communities we design the following minimal, and thus oversimplified – not fully realistic, but illustrative, Dynamic Graphical Model of Pandemic. In a social context where people interact with each other on a regular basis, we assume that the virus spreads in the community (census tract) sufficiently fast, say within five days – which is the estimate for the COVID-19 median incubation period. If an infected person enters a community/neighborhood, but does not stay there, he infects others with some probability. If a single resident of the community becomes infected all other residents are assumed infected as well (instantaneously). Our model is a discrete-time dynamic model where nodes in a network are in one of the three states: **S**usceptible, **I**nfected, or **R**emoved. The nodes represent communities/neighborhood. A contact between an **I**nfected community/node and with another community which **S**usceptible has an assigned probability of disease transmission, or rather turning the **S** state → **I** state. The assigned pairwise probability is symmetric, or rather the probability of transmitting decease from the first community to second one and vice versa are equal to each other, and it depends on the number of factors, for instance on daily traffic between the two communities, current value of *R*_0_, and so on. (The symmetry assumption is plausible, but also not critical for the following derivations. We make this assumption primarily to streamline the notations.) For the time being we assume that the assigned probabilities are known. (We discuss how to set up the assigned probabilities based on the publicly available data in Section IV.)

In what follows we adapt the **SIR** model to the network of census tracts (communities). Consistently with what was described above, the network is represented as a graph, where nodes are tracts and edges, connecting two tracts, have an associated strength of interaction representing the probability for the infection to spread from one node to its neighbor. A seed of the infection is injected initially at random, for example, mimicking an exogenous super-spreader event in the area; examples could include political or religious gatherings. See Figure 3 illustrating dynamics of the cascade model over square grid completed in 8 steps, from random initiation (top left) to the terminal state (bottom right). Color coding of nodes is according to **S**usceptible=blue, **I**nfected=red, **R**emoved=black. Given the starting infection configuration each infected community can infect its graph-neighbor community during the next time step with the probability associated with the edge connecting the two communities. Then the infected community moves into the removed state. The attempt to infect each neighbor is independent of all other neighbors. This creates a cascading spread of the virus across the network. The cascade stops in a finite number of steps thereby generating a random **R**emoved pattern, shown in black in the Figure, while other communities which were never infected, or rather, remain **S**usceptible, are shown in blue. The process known in the Computer Science literature under the name of the (Independent) Cascade Model was introduced and analyzed in a seminal paper by Kempe and company. [74] with many follow-ups [59, 77, 100, 108]. The Cascade Model was set up to study maximal spread of information in a network. In particular, the authors studied the influence maximization in advertising by word-of-mouth: How does one choose nodes in a network to maximize the spread of some information? More generally, Cascade Model was considered as a **diffusion over the network** and its possible application value to modeling and mitigation of the disease spread was mentioned in the Cascade Model literature [59, 77]. This is the simplest model. We discuss more realistic generalizations below.

**FIG. 2:**
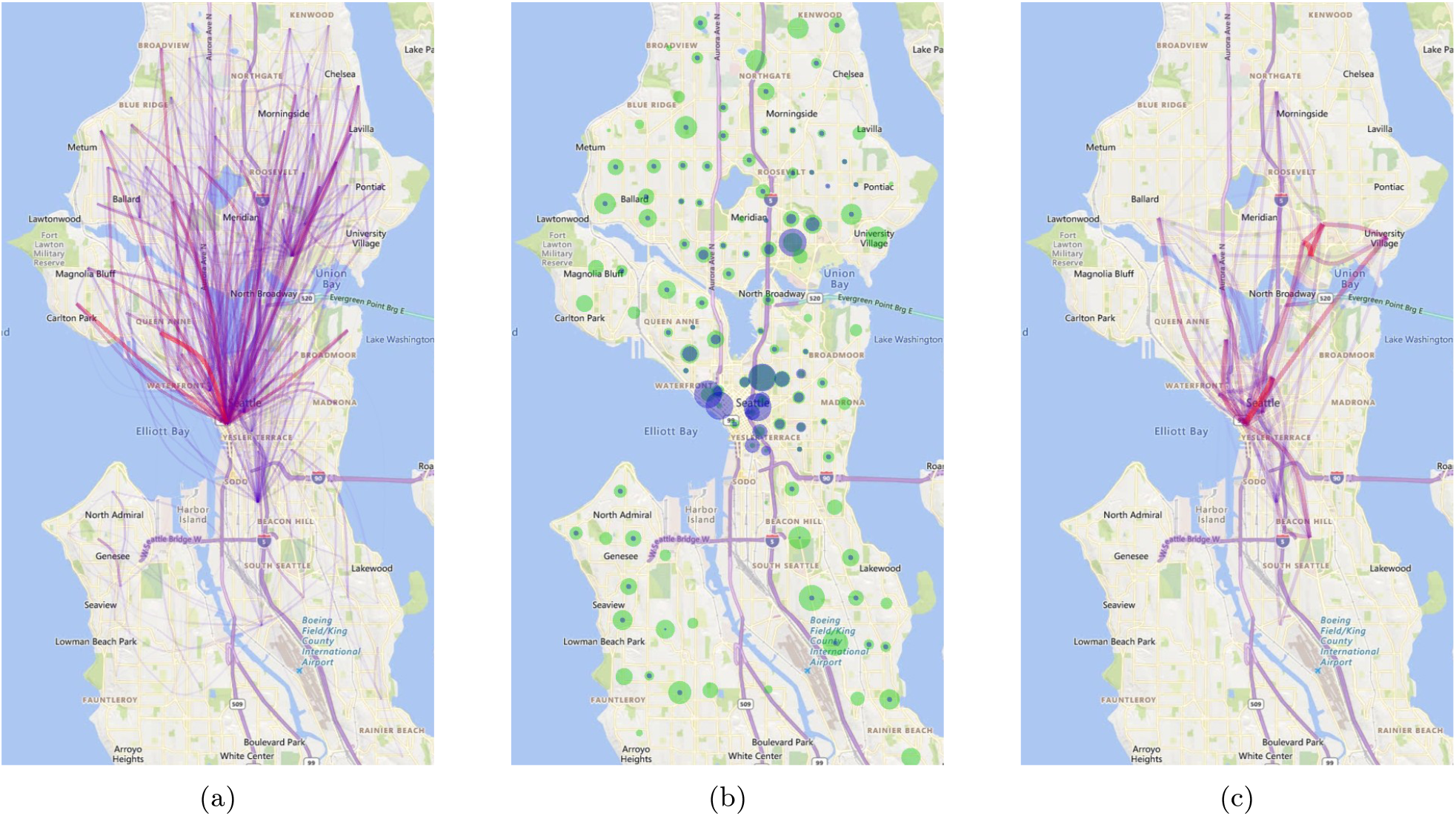
(a): Average travels between census tracts of Seattle. (b): Population density; the green disks represent census tract populations and blue disks represent census tract population densities. (c): Top 20 census tracts with highest travels.

**FIG. 3:**
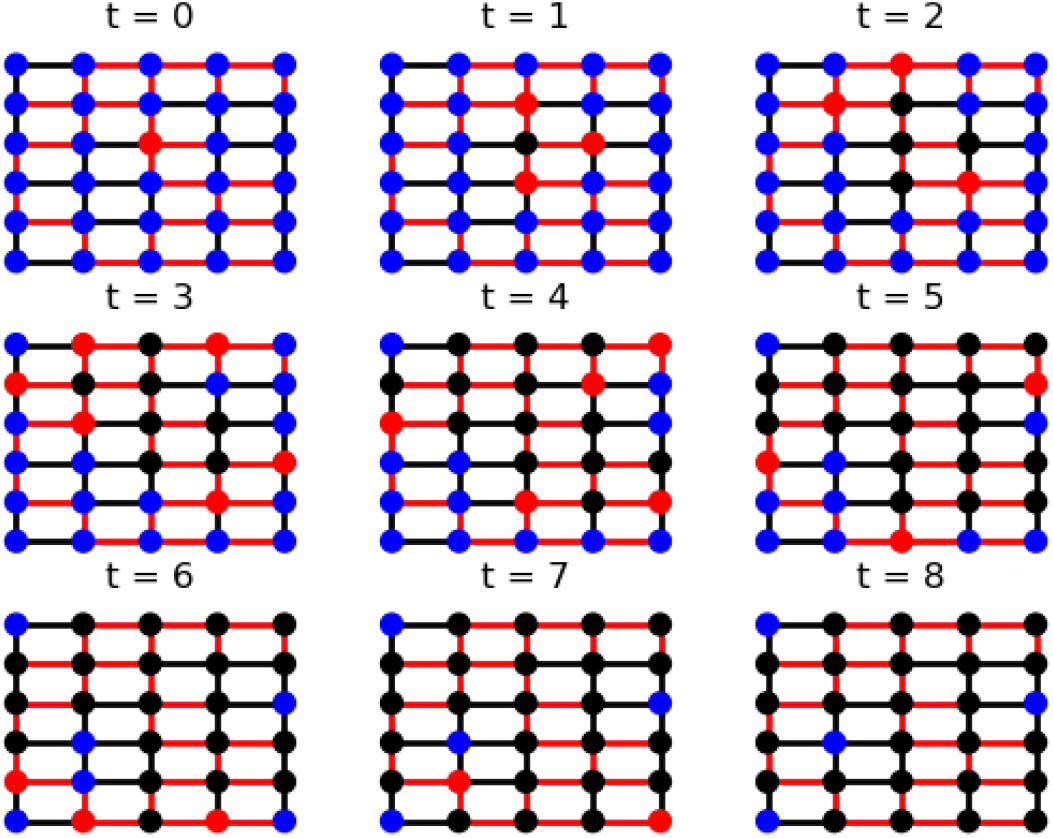
Exemplary sequence of frames illustrating the process of infection spread over square grid, given an initial seed (single node shown red) at the first frame(*t* = 0). Each edge is accessible to transmission in both directions with the same rate. A sample of activated edges is shown in red. (We oversimplify here and consider samples with an edge open in both directions.) A node is (I)nfectious (red) for one time step only, then it becomes (R)emoved (black). The process terminates in 8 steps. (S)usceptable nodes (shown black) were not infected through the process. terminal (R)emoved pattern corresponds to a connected component of the graph formed by activated/red edges containing the initially (I)nfected node.

Notice that the mitigation aspect of the cascade model, which is not the main subject of the manuscript but is discussed briefly in Section VI, requires minimization of influence, that is opposite to what is normally of interest in the context of the information spread.

It was shown in [74], that the terminal **R/S** coloring of the network can be sampled synthetically much faster than through dynamical simulation described above. Specifically, instead of sampling of inter-community infection dynamically, we can introduce auxiliary variables describing activation of edges and pre-sample them efficiently and in parallel, prior to running dynamical simulations. We assign a finite probability to each activated edge indicating if the infection is actually transmitted. This powerful observation results in our principal ability to describe probability of a the **R/S** sample over the network in a static way, completely bypassing dynamics. We simply need to consider sub-graph of the original graph with activated edges (red in the Figures 3,4) and color black (**R**emoved) all nodes connected to the initial seed (initially red nodes) in the sub-graph. This simplification is critical for reducing the Dynamic Graphical Model in terms of its computational complexity even further — to the static Graphical Model described in the following Section.

**FIG. 4:**
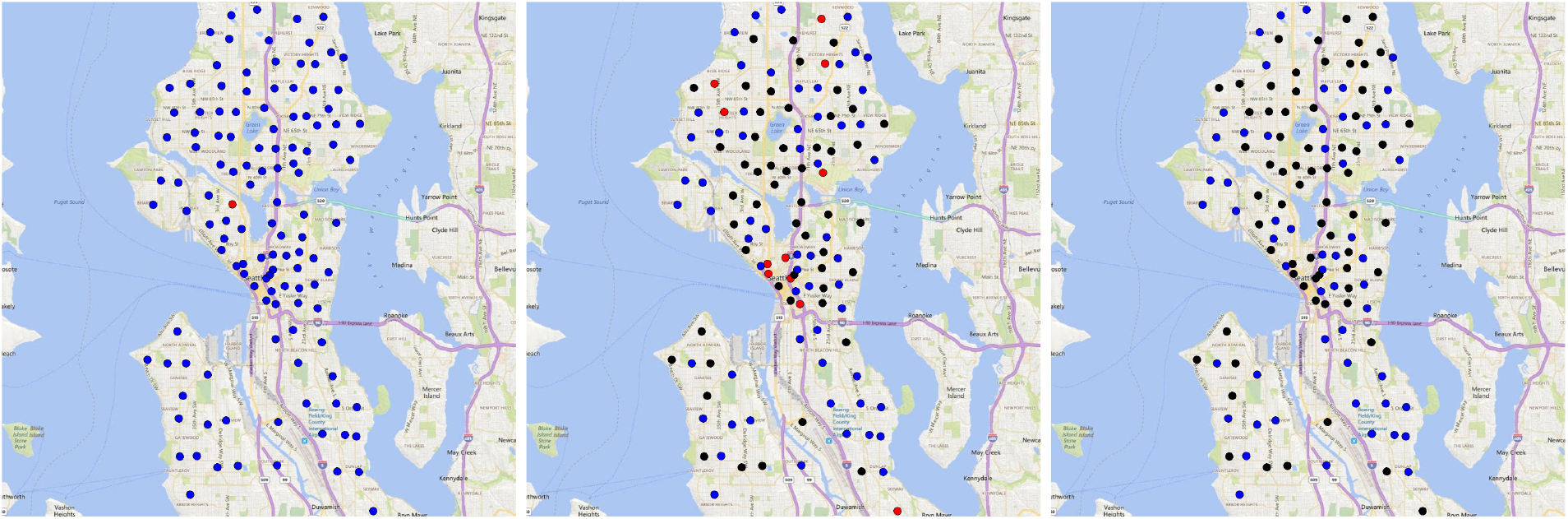
Result of a cascade dynamics on the example of Seattle Network. Color coding is the same as in the square grid Figure above. Initial configuration (left), configuration after 4 steps (middle) and terminal configuration (after 8 steps, right) are shown.

In the remainder of the Section we will provide more detailed information on the cascade model, set notations, and discuss extensions relevant to the Graphical Models of Pandemic.

Consider an undirected graph, 𝒢 = (𝒱, ℰ). If two nodes, *a, b* ∈ 𝒱 are connected by an edge we denote it as {*a, b*} ∈ ℰ, where thus 𝒱 and ℰ are the sets of nodes and edges of the graph. Let us also introduce ℰ_*d*_ for the set of directed edges of the undirected graph and use, (*a, b*), (*b, a*) ∈ ℰ_*d*_, to denote two directed edges correspondent to the same undirected edge, {*a, b*}∈ ℰ, where thus a directed edge, (*a, b*) defines parent, *a*, and child, *b*. We denote, 𝒱_*b*_ = (*a*| (*a, b*) ∈ ℰ_*d*_) ⊂𝒱, the set of parents of the node *b*. Node *a* at the moment of time *t* can be in either of the three states, {*S, I, R*}, **S**usceptible, **I**nfected and **R**emoved, respectively. Introduce *π*(*t*; *a*) denoting the state of the node *a* at the moment of time *t*. The initial infection is seeded at *t* = 0, ***π***(0) = (*π*(0; *a*) ∈ {*S, I*}|*a* ∈ 𝒱), where, ∄*a* ∈ 𝒱 : *π*(0; *a*) = *R*. Note that *π*(*a*, 0) is either *S* or *I*, but could not be *R*. We use *π*(**t**) to denote the vector (*π*(*a, t*), *a* ∈ 𝒱.

Assume that the initial seed, ***π***(0) is given/fixed. Probability of transmitting infection from parent to child is described in terms of the symmetric transmission matrix:

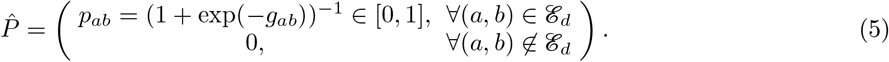

The infection cascade is developed in finite time according to the following dynamic process:

### Definition II.1

(Dynamic Process). *Given a graph* 𝒢 = (𝒱, ℰ) *the rules of the Independent Cascade are as follows:*

- *If a node a* ∈ 𝒱 *is infected at the step, t, it infects its child, b* ∈ 𝒱, *at, t* + 1, *with the probability, p*_*ab*_.
- *Each child (of a parent) is infected independently*.
- *Each node infects its children independently of other nodes*.
- *A node stays infected only for one step and then it is removed and can never be infected again*.

The initial seed, can thus be explained in terms of 𝒱^(*in*)^ = (*a* |*π*(0; *a*) = *I*) *⊂* 𝒱. First obvious observation is that for any seed, 𝒱^(*in*)^, any graph, 𝒢 and any probability matrix, 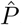, the process terminates, or rather infected nodes disappear, in a finite number of steps. Moreover, it is also obvious that some nodes may remain in the susceptible state by the time of the process termination.

What is less obvious is that the process of infection spread is well defined, in the sense that the PDF of the terminal configuration, ***π***^(*T*)^ = ***π***(*T*), where the time *T* is large enough so that ***π***(*T* + 1) = ***π***(*T*), does not depend on the order of transmission, or rather on the order of the substeps within each step in the algorithm above. This claim of the uniqueness of the outcome, even though in a bit more restrictive form, was made in [74].

Let us pause and clarify what do we mean by the substeps and freedom in the order of transmission within the model. Indeed, the model defined above allows freedom in selecting order of how the currently infected nodes are picked for infection spread to its children. For example, we can pick the infected nodes i.i.d. at random, or we can also pick them according to a predefined order (that is, order which is preset for all nodes of the system.)

Moreover, the authors of [74] have also suggested a construction, consistent with the statement of uniqueness, which allows to replace the dynamic process by an auxiliary static process to describe statistics of 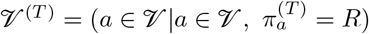.

### Definition II.2

(Static process). *Given the initial seed*, 𝒱^(*in*)^ *⊂*𝒱, *the auxiliary static process of building the outcome*, 𝒱^(*T*)^ (𝒱^(*in*)^), *is defined as follows*

- *Toss a coin for each directed edge*, (*a, b*) ∈ ℰ *of the graph and set the edge to be active (respectively inactive) with the probability, p*_*ab*_ *(respectively* 1*− p*_*ab*_*). We define the set of active edges by* ℰ^(*act*)^. *See Figure 3 for an illustration*.
- *Add all nodes which are infected at t* = 0 *to the list of R-nodes*, 𝒱^(*T*)^ *⊂* 𝒱, *which will be in the removed state by the end of the cascade, or rather set* 𝒱^(*T*)^ *←* 𝒱^(*in*)^.
- *Check any node (one by one) which is not in* 𝒱^(*T*)^ *(initially not in* 𝒱^(*in*)^*) and add it to* 𝒱^(*T*)^ *if it has a path via activated edges to* 𝒱^(*in*)^.

With this definition we arrive at the following statement.

### Theorem II.3

(Static Process Sufficiency). *Given the initial seed*, 𝒱^(*in*)^ *⊂* 𝒱, *statistics of the terminal state of the dynamic process*, ***π***^(*T*)^ (𝒱^(*in*)^), *is unique, or rather it does not depend on the order of the substep execution, and it is equal to the statistics of the outcome of the Static Process II*.*2*.

Two comments are in order. First of all, that averaging (stochasticity) in Theorem II.3 is associated with the activation of the edges. Second, the actual statement of [74] was a bit weaker than of the Theorem II.3: it was made not for the entire PDF, of 𝒱^(*T*)^ and *B*^(*F*)^, but for the equivalence of the average number of the terminal (R)-nodes.

*Proof*. Instead of providing a formal proof, we will only sketch here the main (and very intuitive) idea behind the proof. Analyzing the dynamic process we observe that when an edge is called, because it connects an infected parent to a child, the probability of infection is decided independently on any of the previous steps. Moreover, by construction each edge will be called only once during the discrete time dynamic process. This means that instead of flipping a coin on the fly and deciding on if the edge is going to be active, or rather transmission of the infection from the parent to a child will be taking place, we can as well pre-compute respective probability for an edge to be active initially – before executing the dynamic process.□

## III. ISING MODEL OF PANDEMIC: EFFICIENT INFERENCE

### A. Transition from the Independent Cascade Model to the Ising Model: Binary Representation

In this Subsection we, first, restate the results of the dynamic, cascade model in terms of the binary variables. We will then fix the binary variable, *A*, associated with the initial infection seed, or alternatively introduce a prior distribution for the binary variables, and then discover that when properly regularized the probability distribution of the terminal configuration of *R* nodes, or rather 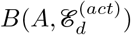, where randomness is due to the random configuration of the activated edges, 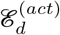, is factorized into a product of graph-local terms according to a Graphical Model (GM) over the original (undirected) graph, 𝒢. In fact, the resulting GM appears to be the pair-wise, attractive, or rather model of the so-called ferromagnetic Ising type.

Assume that the original graph, 𝒢 = (𝒱, ℰ), and the initial infected graph, 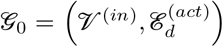, where 𝒱^(*in*)^ *⊆* 𝒱 is the set of initially infected nodes and 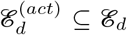 is the set of edges activated according to the static process, are given. Let us also restate the sets in terms of the binary vectors of initially infected nodes and edges:

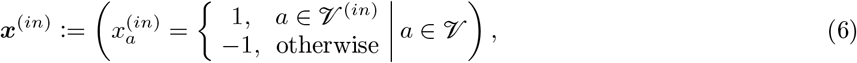

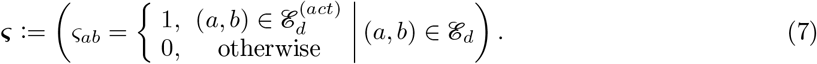

Definition II.2 states the static process in terms of the entire graph-structure. Can one restate it in terms which are graph-local?

To show what we aim to accomplish here (with the locality considerations), consider the following system of inequalities imposed on the vector of the static process output, ***x*** = (*x*_*a*_|*a* ∈ 𝒱):

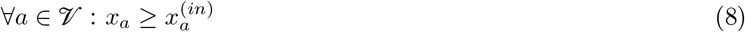

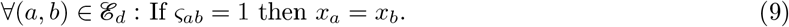

If one starts from the nodes of the initial seed and move to children of the seed nodes via activated edges, and to their children, etc, we observe that at each step of the iterative procedure Eqs. (8,9) are satisfied. Moreover, the equations fix unambiguously all nonzero components of ***x*** corresponding to *a* ∈ 𝒱 with *x*_*a*_ = +1 and achieve this in a graph-local way. That is, each of the conditions (8,9) are stated only in terms of variables associated with a given node and its graph-neighborhood.

However, the Eqs. (8,9) are not sufficiently restrictive to resolve all negative-valued components, *x*_*a*_ = *−*1, of the vector of the static process output.

One way of fixing the remaining freedom/ambiguity in defining the outcome of the static process is to introduce the following optimization

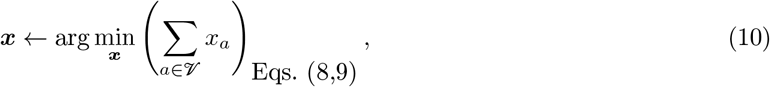

which will effectively set the *−*1 components of ***x*** at all the nodes which are not connected to initially infected nodes via the network of activated links. Therefore, we have just sketched the proof of the following statement.

#### Lemma III.1

(Alternative Definition of the Static Process). *Given a fixed configuration of the initial seed vector*, ***x***^(*in*)^ = {*±*1}^|𝒱|^, *and fixed configuration of the activation vector*, 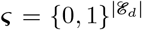, *finding outcome of the static process*, ***x***, *is equivalent to solving the optimization problem (10)*.

At the next step we inject in the description random edge activation according to the following i.i.d. process due to (5):

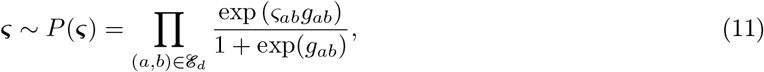

where *ς*_*ab*_ *≠ ς*_*ba*_, or rather, each of the pair of variables is associated with a directed edge. This choice of the *ς*-variables is consistent with the fact that an edge is open for infection the transmission may go in either direction. *g*_*ab*_ in Eq. (11) is the real-valued parameter which measure intensity of direct interaction between nodes *a* and *b. g*_*ab*_ → + *∞* corresponds to the limit when the infection will be transmitted with 100% probability. The opposite extreme of *g*_*ab*_ → −∞ corresponds to complete (direct) isolation of the two nodes. Many microscopic public health parameters, such as current estimations of *R*_0_, traffic closures and other quarantine policies, as well as level of immunization (when vacine is available), influence the *g*-factors directly or indirectly.

#### 1. Fixed Seed of Infection

Let us now assume that the seed of infection is fixed, or rather ***x***^(*in*)^ is given. Then taking into account randomness in the edge activation, expressed by Eq. (11), and expression (10) of the terminal configuration, ***x***, via the initial seed and the set of activated edges we arrive at the following joint probability distribution of ***x*** and ***ς***, conditioned to ***x***^(*in*)^

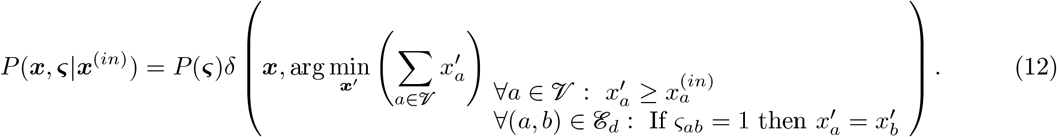

To allow a GM, or rather graph-factorized, representation we introduce a “physical” regularization, which may also be considered as a generalization, of Eq. (12), correspondent to the terminal state of the Independent Cascade model. The regularization is associated with the community (thus independent of other communities) bias towards the S-global, i.e. ***x*** = (*−*1)^|𝒱|^, state:

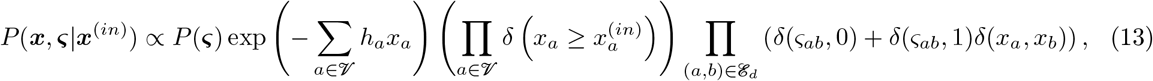

where *δ*(*x*_1_, *x*_2_) is the Kronecker symbol returning unity if *x*_1_ = *x*_2_ and zero otherwise.

If we aim to relate the probability distribution function of the GM (13) to the probability distribution of the terminal state of the independent cascade model given by Eq. (12), we should consider the deterministic, *∀a* ∈ 𝒱 : *h*_*a*_ → + *∞*, limit (also called “zero-temperature” limit in the statistical physics literature). We conjecture however, that the GM (13) also has a sense at finite/bounded, ***h*** := (*β* _*a*_ |*a* ∈ 𝒱).

Indeed, |***h***| *< ∞* may be viewed not only as a regularization vector parameter but also as a parameter expressing additional effects, not represented in the independent cascade model. In particular, *∞ > h*_*a*_ *>* 0 may represent the fact that a node may become infectious due to other, exogenous and independent of other nodes, pathways of the infection arrival at the node. It may also account for a combination of exogenous factors, some leading to increase in the probability of infection, such as frequent violation of the public health policy recommendations (e.g. not wearing masks, violating stay-home-orders), and some to decrease, such as vaccination. Moreover, ***h*** may vary from node to node, e.g. expressing edge, ethnicity, gender or other community-specific biases.

Marginalizing Eq. (13) over ***ς*** and accounting for Eq. (11) we arrive at the following GM formulation for the probability distribution of the terminal state ***x*** conditioned to the initial state, ***x***^(*in*)^:

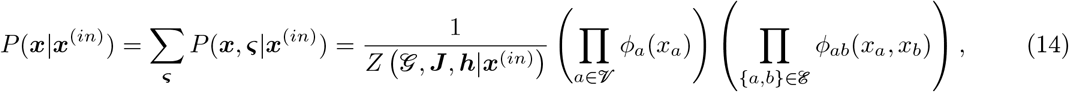

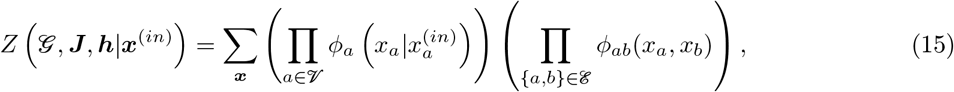

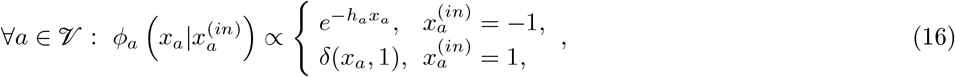

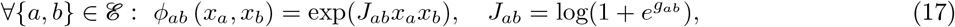

where *Z* (𝒢, ***J***, ***h***|***x***^(*in*)^) is the normalization constant - the so-called partition function of the GM which is conditioned to ***x***^(*in*)^ and also depends on the tuple, (𝒢, ***J***, ***h***), where 𝒢 = (𝒱, ℰ) is the undirected graph expressing the set of neighbors (nodes), 𝒱, and the nodes pair-wise influence via the set of undirectred edges, ℰ; ***J*** = (*J*_*ab*_ |{*a, b*}∈ ℰ), is the |ℰ|-dimensional vector built from positive characteristics, each measuring the strength of the respective pair-wise interactions; and we remind that ***h*** = (*h*_*a*_ |*a* ∈ 𝒱), is the |𝒱| dimensional vector of nodal biases expressing tendency of the respective neighborhood/node to be infected recently or staying immune since the moment of virus injection into the system.

Notice also that transitioning from the deterministic model (12) to the more general (probabilistic) model (13) with finite ***h***, and then resulting (after marginalization) in the Ising model Eq. (14), we also alter the meaning of the pair-wise, ~*δ*(*x*_*a*_, *x*_*b*_), term in the original model (13) and, respectively, the meaning of the pair-wise interaction term ~exp(*x*_*a*_*J*_*ab*_*x*_*b*_) in the Ising model (14). Indeed, consider an edge/pair, {*a, b*}∈ ℰ. In the case of finite 0 *< h*_*a*_, *h*_*b*_ *< ∞*, the pair-wise term represents the tendency for the states of the two nodes to be similar (attract to each other) regardless of the signs of *x*_*a*_ and *x*_*b*_. While “attraction” of *x*_*a*_ = +1 and *x*_*b*_ = +1 is present naturally in the IC model, attraction of the *x*_*a*_ = −1 and *x*_*b*_ = *−*1 states is not represented in the IC model. However, these effects are still realistic in a broader setting, for example representing tendency of the nodes linked to each other to be alike, no mater what is their state. Such effects may express social trends, such as the culture of following the public safety guidance “propagating” from one node to another.

#### 2. Random Seed

We can also consider the case of a random seed ***x***^(*in*)^. Let assume that the seed is injected i.i.d at random according to the probability distribution

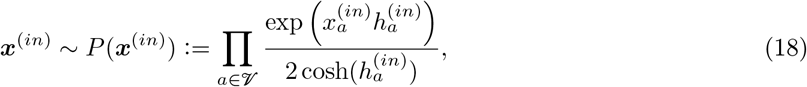

where, 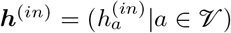 is the vector of the initial infection biases. Then, the *h*-regularized joint PDF of ***x***^(*in*)^, ***x*** and ***ς*** becomes

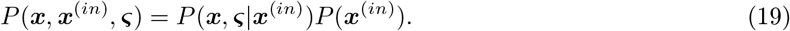

Marginalizing Eq. (19) over ***x***^(*in*)^ and ***ς***, and accounting for Eqs. (13,18) we arrive at the following expression for the *β*-regularized probability distribution of ***x*** in the case of the stochastic initial seed:

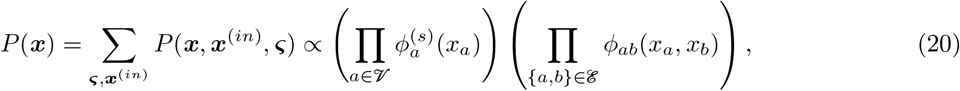

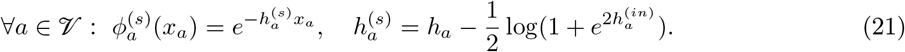

### B. A-Posteriori Measure of the Infection Event

In this Section we discuss a-posteriori marginals and maximal configurations of the Ising model of Pandemic which are of the Public Health interest. We focus on the case of the fixed initial seed, ***x***^(*in*)^, representing injection of virus into the model city and assume that the tuple, (𝒢, ***J***, ***h***), is given.

#### 1. A-Posteriori Marginals

First of all we are interested to compute the Conditioned A-posteriori Level of Infection (CALI) for all the nodes (neighborhoods) of the system (within the city):

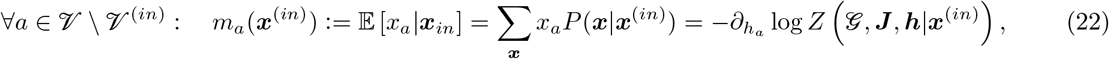

where the Ising model probability distribution, *P* (***x***|***x***^(*in*)^), and the partition function, *Z* (𝒢, ***J***, ***h***|***x***^(*in*)^), both conditioned on ***x***^(*in*)^, were defined in Eq. (14) and Eq. (15) respectively. Four comments are in order: (a) recall that 𝒱^(*in*)^ stands for the subset of nodes where 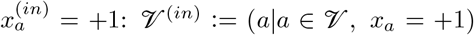 (b) Eq. (22) should also be supplemented (for clarity) by the statement that CALI is equal to +1 at all the nodes where the injection is seeded, i.e. *∀ a* ∈ 𝒱^(*in*)^ : *m*_*a*_(***x***^(*in*)^) = +1); (c) the term commonly used for *m*_*a*_ in the statistical physics literature is the magnetization; (d) Eq. (22) can also be re-stated in terms of the partition function of the bare model (3):

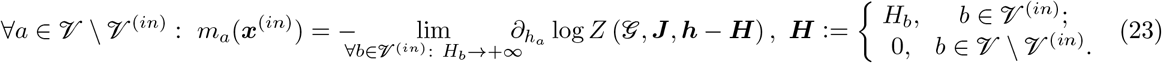

In addition to the CALI we may compute the weighted (over the entire graph) average CALI, which we coin CALI-index:

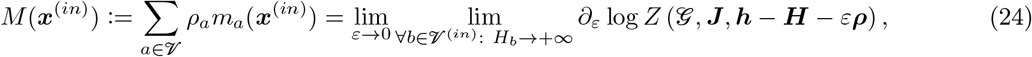

where Σ _*a* ∈ 𝒱_ *ρ*_*a*_ = 1; *ρ*_*a*_*≥* 0 is the normalized population density in the city (neighborhood), *a*; and ***ρ*** := (*ρ*_*a*_ |*a* ∈ 𝒱) is the vector of densities.

CALI itself, defined in Eq. (22), and the CALI index, defined in Eq. (24), both depend on the the initial seed, ***x***^(*in*)^. It is of interest to choose the initial seed, ***x***^(*in*)^, localized at a single node, which we denote, *a*^(*in*)^ ∈ 𝒱 (also, and with a slight abuse of notations, replacing *x*^(*in*)^ by *a*^(*in*)^ in Eqs. (22,24), or rather *m*_*a*_(***x***^(*in*)^) → *m*(*a*^(*in*)^, *a*), and *M* (***x***^(*in*)^) → *M* (*a*^(*in*)^)). This localized choice of the infection’s injection corresponds to the regime when the epidemic is mainly under control locally (within the city) while other external sources of infection are still exist and significant, so that individual travelers/visitors can bring the infection in. Naturally, we study the matrix 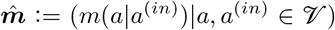, and the vector, ***M*** := (*M* (*a*^(*in*)^)|*a*^*in*^ ∈ 𝒱) and the node-heat-map of ***M***. (Note that 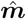 is the skew-symmetric matrix with *∀* ∈ */*+1 on the diagonal, *∀ a* ∈ 𝒱 : *m*(*a, a*) = +1. We show in the Figures the two numbers, *m*(*a, b*) ≠ *m* (*b, a*), associated with the pair of conjugated off-diagonal elements of the matrix, by splitting respective undirected edge, {*a, b*}, in two equal parts and coloring them differently, such that coloring of the part adjusted to the *a*^(*in*)^ node corresponds to *m*(*a* | *a*^(*in*)^).)

The CALI matrix, 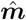, describes the pair-wise response – sensitivity of the terminal result at a node in response to injection of the infection seed at another node.

The CALI-index, ***M***, helps to rank locations according to their overall danger in terms of the terminal consequences of seeding infection at the location. We can also compute the integrated CALI, which we call ALI (not conditioned to the location of the initial infection),

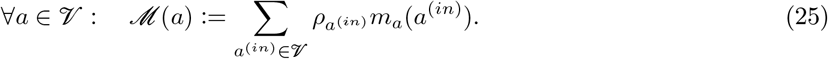

We will also use notation, ***ℳ*** := (*ℳ* (*a*) | *a* ∈ 𝒱), for the ALI vector.

Once ordered in the descending order the ALI vector, ***ℳ***, can be used to set up the vaccination priority, with the node of the maximal ALI, arg max_*a* ∈ 𝒱_ *ℳ* (*a*) to be vaccinated first.

Finally, we introduce the ALI index

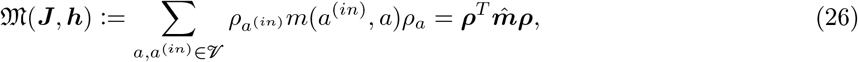

where ***ρ*** = (*ρ*_*a*_ |*a* ∈ 𝒱). The ALI index, 𝔐, which depends on the tuple, (𝒢, ***J***, ***h***), is the ultimate number (scalar) representing impact of statistically homogeneous injection of disease into the system. If control of tuple is available, by non-pharmaceutical or pharmaceutical means, we aim to minimize, the ALI index, 𝔐, over the tuple, possibly subject to constraints, for instance expressing limited availability of the control/mitigation resources. See Section VI F for additional discussions and details.

#### 2. Efficient Computation of the A-posteriori Marginals

Computing explicitly the objects of interests, for instance the partition function of the CALI-vector, CALI-index, ALI-vector and ALI index, all expressed via the partition function of the Ising model and discussed in Section III, is the task of exponential (specifically, so-called #*P*) complexity. In other words, even if the tuple (𝒢, ***J***, ***h***), defining the Ising model is known (defined), to evaluate, for example the ALI index 26, will take computational efforts which is proportional to 2^|^𝒱|, where |𝒱| is the number of nodes in the graph, 𝒢 = (𝒱, ℰ). Therefore, exact computations are infeasible already for general graphs with 20-30 nodes and evaluating the ALI index, as well as other quantities of interest, for cities and counties modeled with tens-to-hundreds nodes (census points) require approximate computations. Luckily the approximate techniques for inference in Graphical Models in general, and in Ising models in particular, are well developed – a number of books is written on the subject [78, 97, 116] and it remains to be a very active area of research with many applications in natural and engineering sciences, in particular most recently in AI and Data Science, driving it (see for instance most recent relevant contributions from one of us [37] and references therein). Some of the techniques are built on special tractable cases, for example GM over tree-like graphs [121], Gaussian GM [71, 91], and planar binary GM [83–85]. The trustworthy approaches originated from the tractable base cases, in particular in relation to the gauge transformation and loop calculus [37, 38], set solid theoretical foundations for variational algorithms, for instance loopy Believe Propagation [116, 121], stochastic (MCMC) algorithms [22, 99] and the algorithms of sequential elimination/marginalization [20, 21, 40, 41, 86], and hybrids [16, 18].

In this manuscript focusing on application of the GM techniques to pandemics (and not on developing the GM methodology) we choose to experiment with Mean Field (MF), Belief Propagation (BP) and Global Bucket Renormalization (GBR) utilizing software developed in [20, 21] and available on github. MF and BP are standard (in GM literature) iterative message-passing algorithms, while GBR is an algorithm of the sequential variable elimination type. Those algorithmic choices are made based on the practical considerations. Specifically, we took advantage of the fact that GBR was adapted in [20, 21] to studies of the Ising models over general graphs and showed superior performance in comparison with other (for instance MCMC, variational, i.e. MF and BP, and hybrid) alternatives.

We will also work below with an extremely simple approximation for partition function and marginals of the Ising models – coined two-state approximation. It consists simply in accounting in the sum over exponentially many states only two contributions, ***x***_+_ = (*x*_*a*_ = +1| *a* ∈ 𝒱), and, 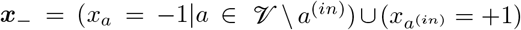, and ignoring all other contributions. We observe that this crude approximation, is surprisingly efficient in some rather wide range of the Ising model parameters considered in the manuscript.

#### 3. Maximum A-Posterior Estimations

The Maximum A-Posterior (MAP) configuration, conditioned to the pattern of the initial infection, ***x***^(*in*)^,

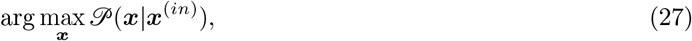

explains the most probable pattern of the infection spread in the terminal state.

It is known that the ferromagnetic Ising model is log-supermodular [43, 116], or rather

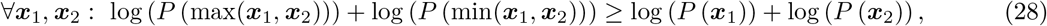

where max and min are evaluated component-wise. The log-supermodularity allows to solve the MAP problem (27) efficiently, by formulating the following tractable LP (also called MAP-LP) over marginal beliefs (proxies for probabilities), see for instance [79, 110, 119]:

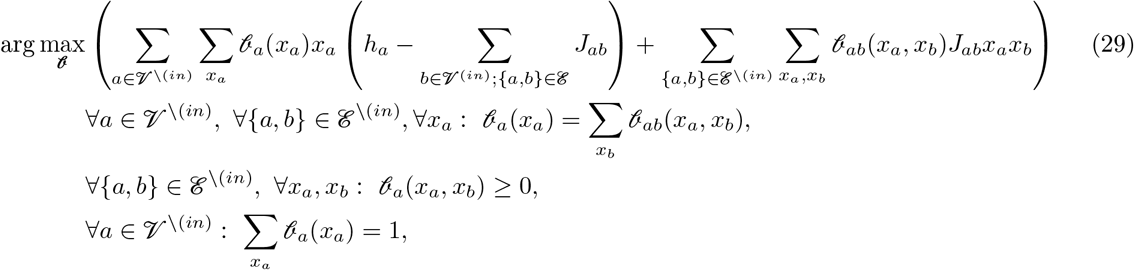

where **𝒷** := (𝒷_*a*_(*x*_*a*_)|*a* ∈ 𝒱^*\* (*in*)^, *x*_*a*_ = *±*1) *⊗* (𝒷_*ab*_(*x*_*a*_, *x*_*b*_)|*a, b* ∈ 𝒱^*\*(*in*)^, {*a, b*} ∈ ℰ^*\*(*in*)^, *x*_*a*_, *x*_*b*_ = *±*)1 ; 𝒱^*\*(*in*)^ is the shortcut notation for 𝒱 *\* 𝒱^(*in*)^ and ℰ^*\*(*in*)^ is the notation for the respective subset of edges, i.e. ({*a, b*}|*∀a, b* ∈ 𝒱^*\*(*in*)^, {*a, b*} ∈ ℰ).

## IV. EXPERIMENTS

The city of Seattle is selected for the case study because relevant data on traveling patterns between 123 census tracts is available as a part of the FLuTE epidemiology simulation library [33] with the additional geographical information coming from the OpenStreetMap [103]. We extract from the data the number of travelers, *N*_*ab*_, moving between each pair of census tracts, *a* and *b*, interpreted as a possible edge, {*a, b*} of the Seattle graph, in five days (remind, tha this is the median time estimation for COVID-19 incubation period, which we thus choose as time step in the independent cascade model). The maximum number of travelers traversing the most significant routes connecting one census tract with others within the five days is *≈* 3, 500. We translate the travel data (number of travelers a week for all edges of ℰ) into the Ising model pair-wise interaction strength, ***J*** = (*J*_*ab*_|{*a, b*} ∈ ℰ), entering Eq. (17), and respectively the value of *g* entering Eq. (11), according to the following, arguably oversimplified formula

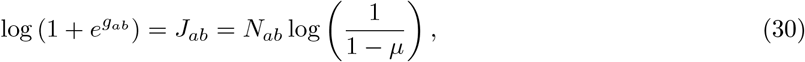

where the left (first) relation is simply the restatement of Eq. (17), while the three-step arguments leading to the right (second) relation are presented below.

1. It is reasonable to assume that a typical visitor meets with *O*(1) infected people during a visit and will touch *O*(1) infected surfaces per visit. Therefore, the probability for such a typical visitor to get an infection during a single visit, call it *µ*^(*t*)^, is not dependent on the size of the visiting community. We estimate that at the pick of a severe pandemic, like COVID-19, *µ*^(*t*)^, is in the [0.01, 0.03] range, which means that it takes the typical person roughly 30-100 trips to the infected community to become infected with high probability. This estimate is roughly consistent with the data reported via accounting for various physical mechanisms of the virus transmission, see for instance [30].
2. However, a typical person does not seem to be a major factor in transmission of COVID-19. As established in [117], based on the extensive phylogenetic studies, small percentage of the infected individuals (as few as 10% and probably less) become the so-called super-spreaders which then infect many more individuals (on the order of 10) in the second round than the value of the averaged rate of transmission, *R*_0_, would suggest. We assume that it is sufficient to get one such a super-spreader returning infected from a trip to a neighboring community to turn the entire community from **S**usceptable to **R**emoved in a few weeks after the travel. (Note in passing that all consecutive, beyond secondary, transmissions originating from the super-spreader) are not modeled within the coarse-grained Ising model, but simply assumed.) Reconciling this (10%) estimation with the one from (1) above, we conclude that the probability that a visitor will bring an infection home in the result of a single trip and also that the visitor is a super-spreader becomes, *µ* = *µ*^(*t*)^ * 0.1 ∈ [0.001, 0.003].
3. Finally, the probability that at least one of the *N*_*ab*_ (independent) visits in five days (remind that the 5 days is the ICM time step) results in the super-spreader mechanism, discussed in (2), becomes 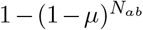. This estimate equated with 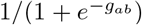, also consistently with the definition of *g*_*ab*_ in Eq. (11), results in the right hand side part of the Eq. (30).

We set the value of the community/node local bias in the Ising model “Seattle” experiments to be the same for all the nodes, *∀ a* ∈ 𝒱 : *h*_*a*_ = *h*. (We remind that *h* introduces the “regularization” bias towards the **S**usceptable (−1) state, and it can thus be called a healthy bias.) We choose to experiment with different positive (bias towards healthy) *h*, ranging from 0.05 to 5. (We remind that predictions of the Ising Model become asymptotically equivalent to predictions of the terminal state in the Independent Cascade model only in the limit of *h* → ∞, when *J*, and respectively *µ* are kept constant. On the other hand, and as we have argued above, considering regime(s) of relative small *h*, when compared with *J*, has a sense have a sense in the broader context accounting for additional effects not contributing the ICM, such as vaccination, enforcing public safety rules and connected communities aligning with eaach other in terms of following the public safety rules.)

As mentioned above making inference in GM, e.g. computing CALI defined in Eq. (22), is a problem of complexity which grows exponentially with the system size. It means, in particular, that making computations over the 123-nodes-large GM of Seattle we need to rely on approximations/heuristics. We use the state of the art heuristics, which we also calibrate on a scale down GM of Seattle. Specifically, we have created 10-nodes-large and 20-nodes-large models of Seattle by aggregating census tracts into 10 and 20 geographical areas, and then constructing the interaction matrix between the nodes/areas in the same way as described above for the case of the 123-nodes-large GM of Seattle.

The results of our experiments with the 10-nodes-large model of Seattle are shown in Fig. 5, reporting only results of exact (within the model) computations. This scale down experiment returned a number of interesting observations:

**FIG. 5:**
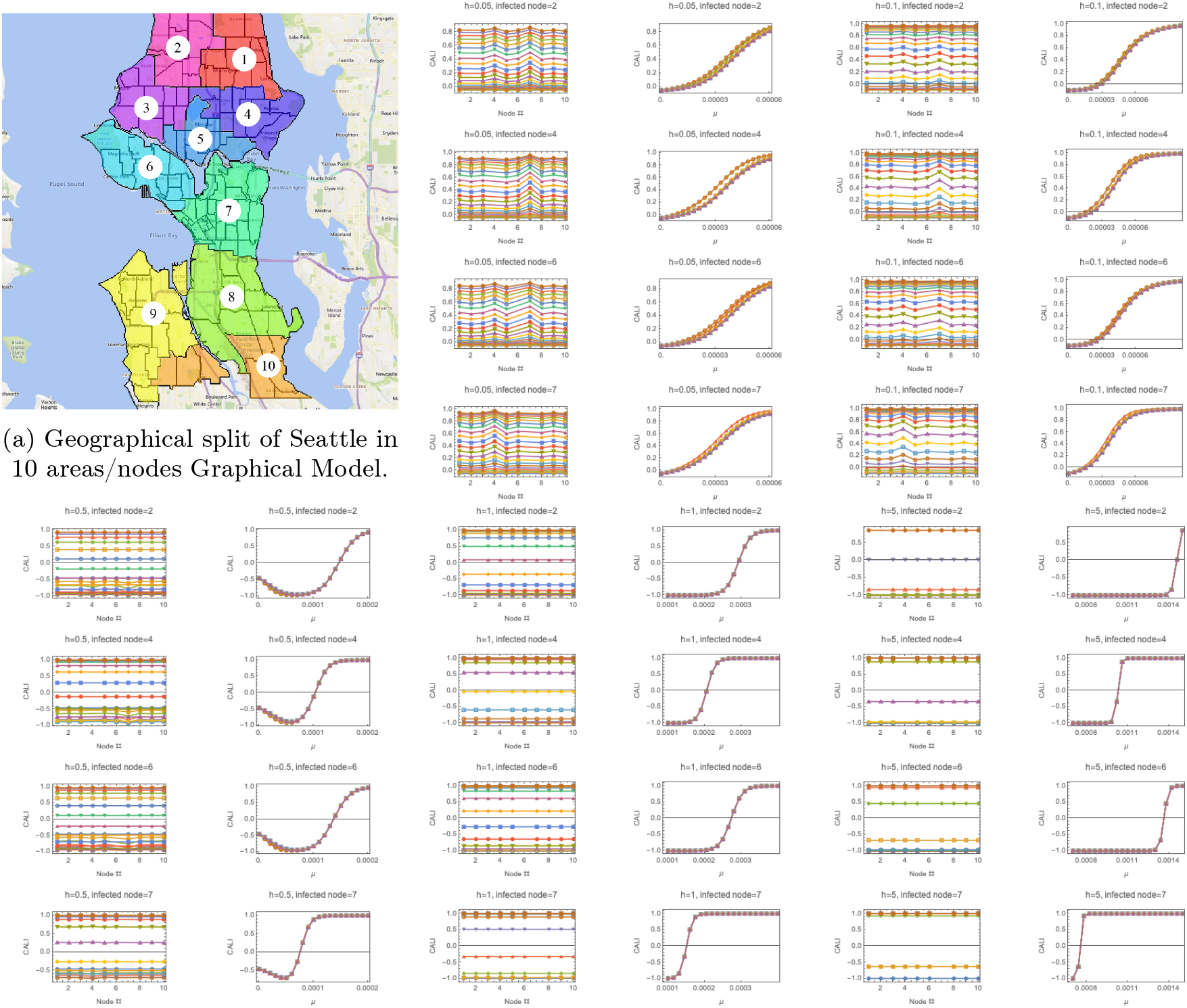
CALI evaluated (exact inference) over the 10 nodes GM of Seattle. Each pair of subfigures (CALI vs node id and CALI vs *µ*) corresponds to one set of experiments for fixed (and uniform) bias *h* and showing results for different values of the interaction factor and different initial seed of infection (infected nodes). See discussions in the text.

- We observe cross-over with increase in *µ* from not infected (large *h*), or largely not infected (small *h*), state to fully infected state with increase in *µ*. This behavior is expected. What was, however, not expected is that this cross-over is not always monotonic. Specifically, in the regime of moderate *h* and sufficiently small *µ*, increase in *µ* leads to decrease in CALI. This, somehow surprising, observation is linked to the attractive nature of the interaction which makes alignment of the nodes “communicating” with each other stronger more preferable. Therefore, in this situation increase of the traffic (that is increase in *µ*) has a very positive effect. Such effect may be attributed, in particular, to “export” of positive habits from one community to another, e.g. practice of wearing mask and practice of keeping social distance. This extremely positive effect of the traffic increase may be extreme, e.g. as seen in the *h* = 0.5 regime at *µ ≈*0.00005 (see Fig. 5) where CALI are very close to *−*1 for all nodes (but the initially infected one), i.e. all nodes are almost infection free. Notice, however, that further increase of the traffic (increase in *µ*) leads to a rather sharp transition to the undesirable (all-infected) state.
- In almost all considered regimes CALI was either exactly uniform (within the computational precision) or it was close to uniform. This observation is consistent with the statement that, of the exponentially many possible global states of infection (2^9^ in this model), the two states – all-infected and all-infection-free (but one initially infected) – are realized with much higher probability than other states. Moreover, we observe a “phase transition”, or more accurately (as strictly speaking “phase transitions” attribute to the thermodynamic limit) strong cross-over/threshold behavior, with increase in *µ* in the regime of sufficiently large bias, *h*. As discussed below, the uniform expectation for CALI may become instrumental for building very efficient inference algorithm even for much larger (and more realistic) Graphical Models of Pandemic.
- The aforementioned cross-over with increase in the inter-community interaction strength (increase in *µ*) is sensitive to the initially infected node. Specifically this (rather sharp) cross-over may occur at different values of *µ* depending on the node infected. See, for example, sub-figures in Fig. 5 correspondent to *h* = 5: if the infection is injected at the node #7 (the Capitol Hill area) this undesirable cross-over takes place at smaller *µ, µ ≈* 0.0007, than if the infection starts at the node #2 (North of the Ballard area), *µ ≈* 0.0014. This observation suggests, in particular, that more detailed monitoring of the epidemiological situation may be needed in the areas leading to prediction of the cross-over at a lower interaction rate.

The results of our experiments with the 20-nodes model of Seattle are shown in Fig. 6. We use this intermediate size model to validate different approximations. Specifically, we juxtapose the exact result against GBR18 – Global Bucket Renormalization with the ibound parameter set to 18 (ibound=20 would correspond to the exact), Mean Field (MF) and Belief Propagation (BP) approximations. (See [20] for details.) We also compare the exact result with the two-mode approximation, accounting for only two extreme states in the Ising model. (We remind that the two mode approximation is expected to be valid when variability of CALI at different observed nodes is small.) Experiments with the 20-nodes model confirm qualitative trends observed in the 10-node experiments, and in addition yield the following observations (and conclusions)

**FIG. 6:**
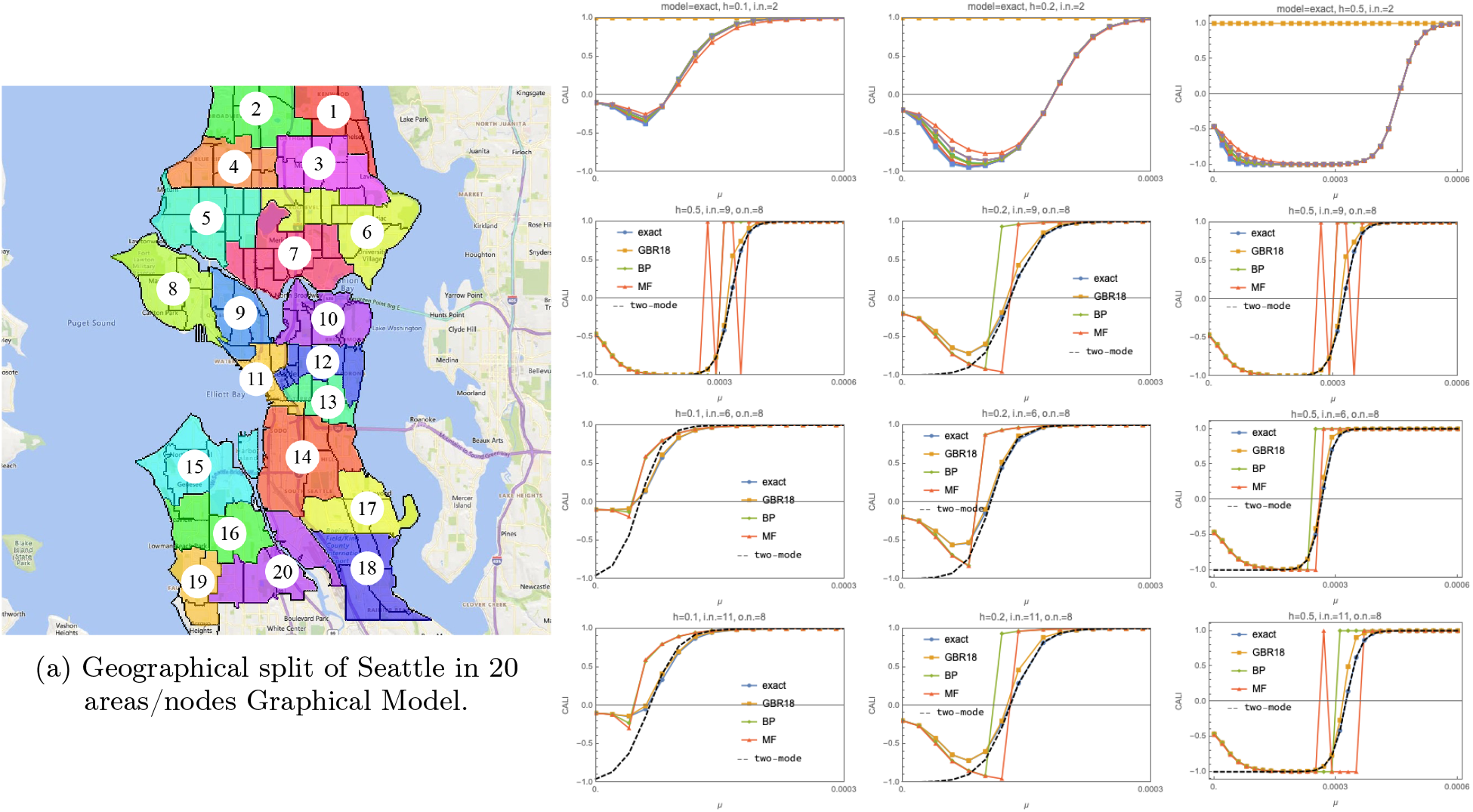
CALI evaluated over the 20 nodes GM of Seattle, with the split (into nodes) shown on the left. Each column of the sub-plots corresponds to three different values of local bias (from smaller on the left to larger on the right). Top row shows CALI at all the observed nodes (o.n.) for the case when the initial infection is seeded at the node #2 calculated exactly (model=exact). Three bottom rows show CALI, computed with different methods (exact, GBR18,BP and MF – see discussion in the text), at the observed node #8, local bias correspondent to the respective column and injected at various nodes (#9, #6 and #11 at the second, third and fourth rows, respectively).

- The two-mode approximation is always perfect (matches exact) at sufficiently large values of the pairwise factor, *µ*. Moreover, the interval of the good-to-perfect matching starts at the values of *µ*, where CALI is sufficiently close to *−*1 (observed nodes are largely not infected, in average).
- The two-mode approximation deviates from the exact (it underestimates infection) at small values of *µ*, where, however, BP and MF are performing well (i.e. are sufficiently close to the exact result).
- There exists a range of intermediate *µ* where MF and BP overlap with the two-mode solution – all providing a quality approximation to the exact result. Transition of CALI with increase of *µ* from negative to positive values is not reproduced well by BP and MF.
- GBR18 follows the exact all the way showing the largest deviation in the transient regime, where on the other hand the two-mode solution shown a better matching to exact.

As mentioned above exact computations of CALI and other derived characteristics for the full 123-node model of Seattle is not feasible. However, inspired by excellent performance of the two-mode approximation validated on the smaller (coarser) models, we carry the approximation on to the 123-node model. Specifically, we computed dependence of the CALI-index, defined in Eqs. (24), on the position of the initial seed and on the strength of the pair-wise interaction factor, *µ*. The results are shown in Fig. 7. We observe a rather strong sensitivity of the overall intensity of how the infection spread over the entire city (this is the description of the CALI-index functionality) on the initial seed location and on the value of *µ*. At sufficiently small values of *µ*, correspondent (for example) to overall reduction in the overall traffic of agents over the city, the infection succeeds (i.e. it spreads to higher percentage of census tracts in the city) only if seeded in the higher-traffic locations (left panel in Fig. 7). On the other hand if the inter-city traffic of agents is enhanced (larger values of *µ*) there are only a few census tracts, which are more isolated then others from the rest of the city, do not lead to a city-global spread of the infection if it is seeded at these locations. In these illustrative experiments, the particular values of the homogeneous bias, *h*, and the range of values of the pair-wise interaction factor, *µ*, were selected based on the arguments given in the beginning of the Section. Additional experiments, e.g. with other approximate methods and exploring wider range of parameters and additional complication (such as census tract local bias, *h*_*a*_, in-homogeneous, i.e. dependent on *a*) are in progress.

**FIG. 7:**
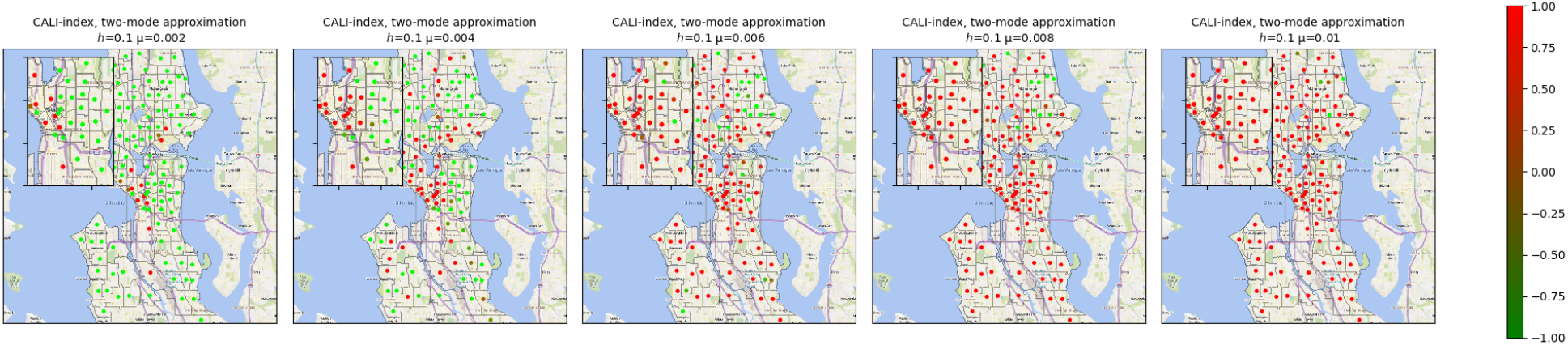
CALI-index, computed within the two-mode approximation, is shown for the full (123-nodes) model of Seattle. It shows dependence of the overall (integrated over entire city) level of infection dependent on the position on the initial seed. Sub-figure on the left, correspondent to reduced pair-wise interaction factor, *µ*, shows that the infection spread succeed only if seeded at a few location visited often by non-residents. Increase in *µ* (sub-figures on the right) results in a much stronger spread of infection. See explanations in the text for further details.

**FIG. 8:**
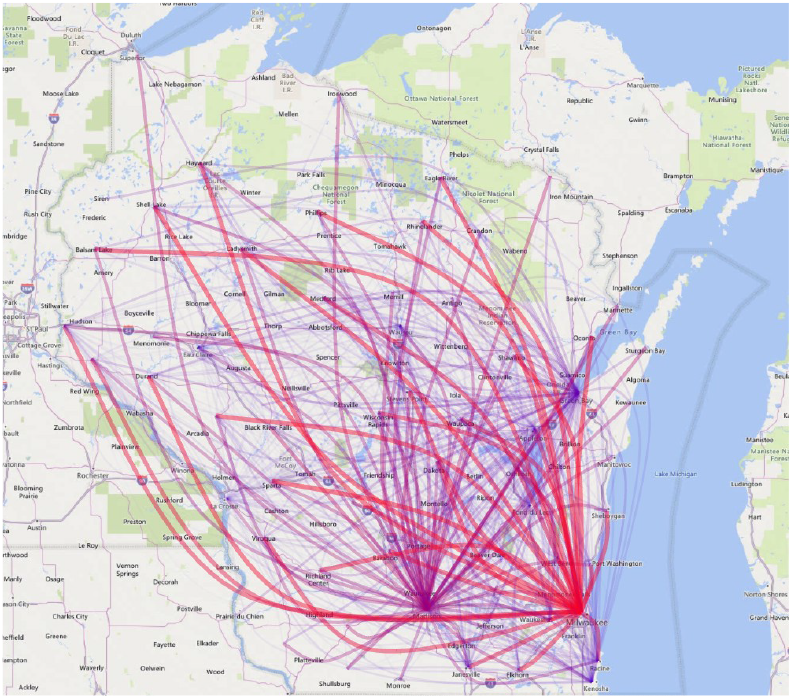
Travels among counties in Wisconsin.

**FIG. 9:**
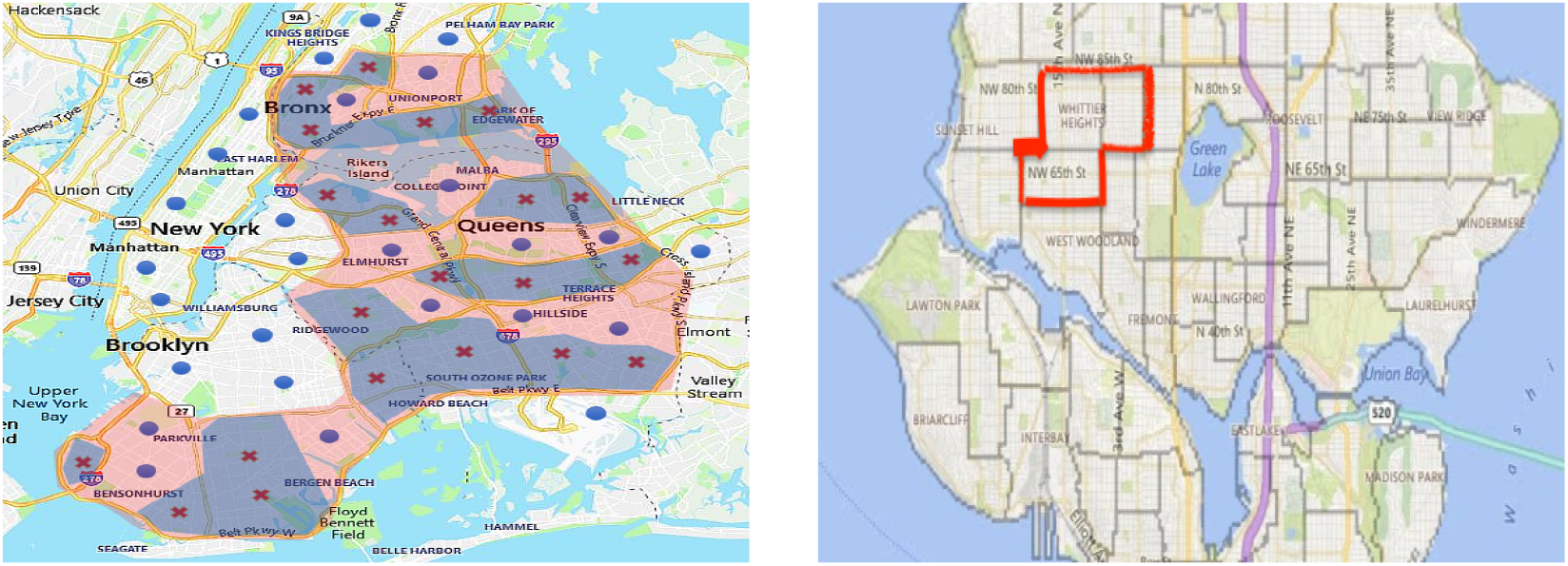
Schematic illustrations of the quarantine decisions. Left Panel: Example of NYC. Blue shedding mark areas with higher transmission (crosses) and low transmission (disks) optimally, or rather with small number of outliers. By contrast the pink separation is more compact even though not outlier optimal. Right Panel: Example of the Enclosed region in Seattle. The boundaries are census tracts.

Let us stress that the choice of the interaction vector, ***J***, and of the local bias vector, ***h***, just described is illustrative and not yet fully realistic. It is introduced in the manuscript for the sole purpose of illustrating methodology. See further discussion of how we plan to make the reduced model more realistic and practical in Section VI.

## V. CONCLUSIONS

This manuscript is the first step in the authors attempt to develop methodology for the data → ABMs → GMs pipeline which aims at improving our modeling, predicting and ultimately mitigating epidemics and pandemics. Specifically the paper is focused on:

- Motivating the use of the Graphical Models as a reduced modeling tool to describe the pandemic;
- Adapting Dynamic Graphical Models (DGM) of the so-called Independent Cascade Model (ICM) type, developed in Computer Science to describe diffusion of influence over the network, to the spread of the infection over the network of communities (e.g. census points). The stochastic dynamics is initiated by an exogenous injection of the infection at a node (or set of nodes), where the network represents a city or a state, and terminates in a sample, where each node (community) is in either of the two states, **R**emoved – with a significant penetration of the fresh infection during the span on time between the initial and terminal configurations, or **S**usceptable.
- Showing that the terminal samples in a naturally regularized DGM of the ICM type are described by an static GM of the attractive Ising Model type, where pair-wise interactions represent the strength of daily exchange of visitors between nodes and the singleton (node-local) term represent initial injection and also a counter-balancing bias of the community towards the **S**usceptable state.
- Building an exemplary and synthetic Graphical Model (of the publicly available travel and census track data) for the city of Seattle and using it to illustrate the power of inference/predicition one can make for the model. Exact inference in the Ising Model of pandemic is sharp-P-complete (that is of the complexity which is exponential in the number of nodes), therefore we utilize efficient heuristics to derive the results.
- Experimenting with the tasks of inference over the GM of pandemics in Seattle has led to the following conclusions.
  – We show that CALI and CALI-index, defined respectively in Eq. (22) and Eq. (24), are useful characteristics for tracking the spread of epidemics and pandemics. CALI is describing increase in the level of infection at a node of the system graph (census tract) to the initial infection seeded at another node. CALI-index is derived from CALI by integration – it shows overall infection spread depending on the initial seed. We experimented with different values of the local bias, *h*, and intensity of the pair-wise interaction, *µ*, to study how CALI and CALI-index change.
  – We reason the choice of the values picked for the experiments, however we would also like to remind that calibration of the parameters based on real data is one most important aspect of the team efforts which is yet to be developed to make the results fully practical. Therefore, the main focus of the study reported in the manuscript, was on testing and developing the inference methodology – assuming that the GM is stated (i.e. learned/trained/calibrated based on data and, possibly via synthetic generation of data with ABMs – as discussed above). In this regards, we have demonstrated that approximate inference methods developed in the GM community can be used for the problem at hand. we consider Belief Propagation (BP), Mean Field (MF) and Global Bucket Renormalization (GBR) approximations (see [20] and references therein for additional details) and also introduce, based on our observations an extremely efficient two-mode model. All the approximate models were calibrated on 10-node and 20-node coarse-grained versions of the model of Seattle, where comparison with exact computations are feasible. Given the disclaimers above, we made a number of useful observations.
  – We observe that the cross-over from not infected to fully infected state with increase in *µ*, may be not monotonic. In the regime of moderate local bias, *h*, and sufficiently small pair-wise interaction the Ising model suggests that increase of the inter-community interaction may lead to decrease in the overall infection. We attribute this effect, in particular, to the fact that increase in the communications may also mean alignments of positive protection habits among neighboring census tracts.
  – When the interaction is sufficiently large, we observe strong mutual alignment of different census tracts. Remarkably, the alignment is observed still in the regime is not yet fully spread. This observation us useful in two ways. First, it allowed us to design an extremely simple two mode approximation – which allowed to extend efficient computations of CALI and CALI-index to the full model of Seattle. (We also expect it to be practical in analysis of a much larger – national scale – models.) Second, it shows that strong alignment of still minor infection spread among sub-communities can be used as an early indicator of the inevitable further development of the infection spread.
  – We discover a very significant sensitivity of the infection spread to where the initial infection is seeded (e.g. by a super-spreader or due to a super-spreader event). The census tracts are not equal and infecting some may lead to immediate spread to the entire system, while infecting others will not lead to major global spread of the infection under the same conditions. In other words, we show the tools we develop are helpful for quantitative monitoring of the current epidemiological situation with a sufficiently detailed spatial resolution/differentiation.

## VI. PATH FORWARD

Reduced Graphical Models introduced and discussed above show a strong potential for monitoring and control of current and future endemics and pandemics. However, much more work is required to turn the models into a practical tool which is the ultimate goal of this manuscript’s authors. This Section is devoted to sketching tasks which need to be resolved to achieve the goal. The material discussed in this Section should be considered as preliminary, provocative and also inviting others, especially with complementary expertise and approaches then ours, to criticize and collaborate.

### A. Working with the data – from methodology to practice

**Task #1: Preparing Ground Truth Data** for our inference, learning and mitigation efforts described in the following requires working with the ABMs. Our discussion of the ABM models so far was limited to the open source FLUTE platform: [33]. We will continue working with FLUTE but also plan to use other ABMs, e.g. from the groups around the World contributing the MIDAS repository of models [9].

New data sources, such as these publicly available via the SafeGraph Data Consortium [10], in particular, the anonymized population movement dataset [11] representing 45 million smartphone devices that have opted into location tracking [102], can not be integrated in the Graphical Models directly because the data comes in the agent-based format, that is the format used in the microscopic ABMs. We ought to account for the new data sources integrating these into ABMs which are then used to generate coarser data, represented via synthetic statistical samples then fed into Learning of the Graphical Model – see Section VI E.

In addition to new tracing app data available in the era of COVID-19, we ought to use public health expertise to calibrate the choice of many parameters incorporated into the ABMs. In addition to adjusting the basic reproduction rate, *R*_0_, to its current and possibly location dependent value, for instance in conjunction with the up-to-date viral/biological estimates [115], we have to account for parameters controlling, in particular (a) transmission probabilities of the asymptomatic and symptomatic: carriers [61]; (b) contact rates associated with travel, employment, schools, and so on.; severity and fatality ratio [2]; pre-existing immunity [111]; rates of the false positive and false negative testing [15]; uncertainty in the duration of the incubation period [26], and other. Additionally, the epidemiological and demographic data should be merged with GIS data to represent different layers. Epidemiological data through time is to be used to represent the trend of disease spread. At each time-stamp, the past can be used to predict future trend and the future data can be used to validate the results.

### B. Extend Cascade Models

**Task #2: Restyle the Cascade Model** to model spatially coarse-grained and dynamic, thus spatiotemporal, spread of Pandemic. We ought to consider realistic generalizations such as: (a) Richer state space of an individual community, discrete with alphabet larger than three (**S**,**I**,**R**) or continuous; (b) More flexible, and possibly continuous time steps. Focus, first, on statistical properties of the terminal state of cascade over the entire network, then analyzing statistics of the dynamical transient.

### C. Approximate Inference Algorithms

**Task #3: Approximate Inference**. There exist three principally different approaches to approximate inference in general GM [116]: variable elimination [20, 21], variational [19, 36, 38], and stochastic (MCMC– Markov Chain Monte Carlo) [17]. (Early work of one of the authors is cited. See also references therein.)

a. We develop variable elimination approaches suitable for the Ising Model. We intend to adapt the family of approaches developed within the team, coined mini-bucket renormalization [20, 21]. The adaptation will consist, in particular, in taking advantage of the order in which the nodes/communities are eliminated and also adjusting approximation (renormalization) to allow reduction to a GM which allows exact inference or easier approximation. We have started to explore the approach in [35]. In the pilot work we use the algorithms developed in [20, 21] to compute the partition function of a GM. Then, we can obtain marginal probabilities by computing partition functions of modified copies of the original GM (each copy with a single node removed). This will allow us to determine the probability that each census tract contracts the virus over a certain time frame given an initially infected tract. Testing the influence of initially infected tracts can inform public policy on how individuals should quarantine to mitigate the spread of the virus.
b. If graph 𝒢 is planar and the vector of the single-community bias, ***h***, in Eq. (21) is zero, resolving the inference problems exactly can be done efficiently, or rather, in the *O*(*n*^3*/*2^) steps [29], where *n* is the number of nodes/communities. As shown recently by the team in [83–85], the exact approach extends to quasi-planar obtained by gluing successively graph components with *O*(1) vertices along subsets of at most three vertices in a tree-like fashion. Some of the GIS-induced community graphs, or their properly coarse-grained graphs, may show this (potentially desirable) quasi-planar structure or structure, which is somehow close to the quasi-planar graphs, however with ***h*** *≠* 0. We plan to take advantage of this observation and extend the approach of [57] building an efficient upper bound approximations to the partition function considering a convex mixture of tractable quasi-planar models.
c. The log-supermodularity of the attractive (ferromagnetic) Ising model (28) allows not only to compute efficiently MAP-values of interest discussed in Section III B 3, but also suggest a pth to building the so-called “low-temperature” perturbative analysis for estimating the respective partition function and marginals of the Ising Models of Pandemic in the “low-temperature” regime where pair-wise interactions, *J*, are sufficiently large [64].
d. We attempt to synthesize the aforementioned approaches with our earlier work on approximate inference through loop counting [58], efficient MCMC sampling [17], and variational inference [19]. The use of the loop-series approach, originating from [36, 38], based of the so-called Belief Propagation method of the variational inference, also combined with MCMC, is well justified in the case of the attractive Ising model because of the strong mixing properties of the models associated with their Fully Polynomial Randomized Approximation Scheme status [70].

### D. Soft Ising Model of Pandemic: Efficient Inference

The Ising Model of Pandemic explained above is the simplest relevant Graphical Model with a geographical (GIS) and transportation degrees of freedom accounted for. Many generalizations are possible, in particular with richer phase space extending from two states (**S** and **R** per node at the terminal stage, and **S, I, R** in the dynamic case) to as many states as deemed needed for the proper level of resolution.

Natural continuous valued, therefore **soft** generalization of the Ising model (14), which we coin Soft-Ising (S-Ising), becomes

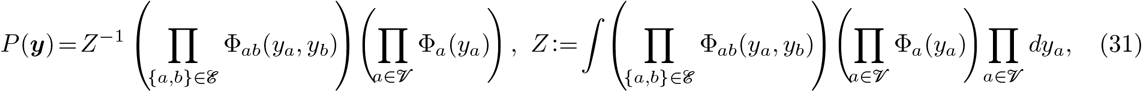

where the terminal state of the system achieved after a cascade transient is modeled with the continuous nodal variables, *∀ a* ∈ 𝒱 : *y*_*a*_ ∈ ℝ, and the extremes, *y*_*a*_ = *−∞* and *y*_*a*_ = + *∞*, correspond to the fully **S**usceptable (cleared from infection) and fully **R**emoved (extremely infectious) states of a node/community, respectively. Sensible exemplary and simple (Gaussian) form of the single-node and pair-wise potentials to consider is

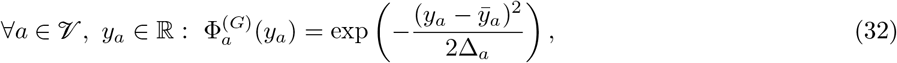

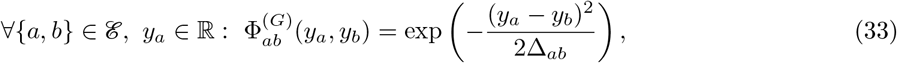

where meaning of the parameters is as follows: 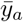 *a*nd Δ_*a*_ are mean and variance of the “prior” (to the initial injection) probability distribution of the state of pandemic at the node/community, *a*; and Δ_*ab*_ is the variance of the mismatch of the pair {*a, b*}of the neighboring communities achieved at the terminal state. Then the exogenous initial injection can be modeled by setting the means, 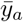, at the initially infected nodes, *a* ∈ 𝒱^(*in*)^, to be significantly smaller (shifted towards *−∞*) than respective values at the initially not infected nodes.

It may also have a sense to consider “beyond Gaussian” statistics. We may need to account for significant intermittency in the pair-wise term (33) modeling effects of non-Gaussian nature of transmission of the infection between communities, for instance analyzing effect of the extended (exponential) tails represented by the Laplace version of the pair-wise distribution:

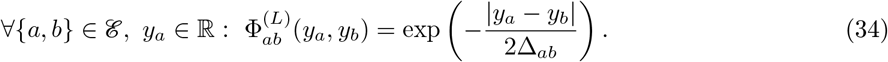

In the case when interaction between neighboring communities is multi-mode it is logical to consider pair-wise potentials in the form of the Gaussian mixture

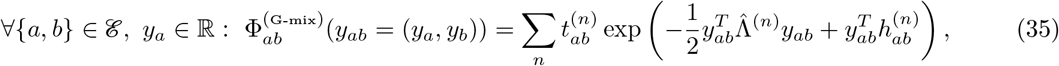

where the positive scalar, 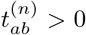, the (2 *×* 2) positive-definite matrix, 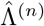, and the 2-dimensional vector,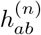, are parameters describing *n*-th mode of the interaction.

Another, and now node-local, modification of our basic Gaussian version of the S-model may consist in replacing Eq. (32) by the Gaussian mixture

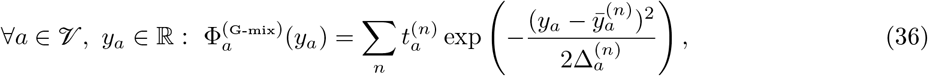

where *n* = 1, 2, … enumerates significant modes of the prior community bias, each characterized by the node-dependent parameters, 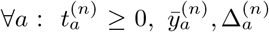, representing split within the communities in groups with different level of immunity, type of vaccination, age, gender, ethnicity, general level of health, and so on.

Our last (and probably most realistic) example in the new family of the S-Ising Models will be mixed GMs [34, 65, 82] combining elements of the Gaussian models (continuous variables) and the Ising models (discrete variables). For instance, we may consider the basic model (31) with the pair-wise potentials removed 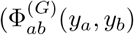 substituted by unity) and the Gaussian nodal potentials (32) with 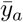 and Δ_*a*_ corrected to be dependent on the auxiliary binary variable, *x*_*a*_ ± 1, via, 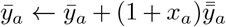, where *x*_*a*_ is random according to the Ising Model of Pandemic discussed in the main part of the text. In this mixed model the 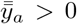 corrections to the local a-priori biases represent increase of the **S**/**R** proportion at the node, if it acquires a “fresh” random infection through a system-global sample of ***x*** with *x*_*a*_ = +1.

**Task #4: Inference in the S-Ising Model of Pandemic**

a. Derive static S-Ising Model from a properly generalized, or rather, continuous space, cascade model. This derivation should help to relate parameters in the continuous space cascade model to factor functions in the C-Ising model (31). We envision adapting the cascade model and introducing continuous valued pair-wise transition probability – when graph-neighbor communities *a* and *b*, which are in the continuous valued states of infection, *y*_*a*_, and, *y*_*b*_, in the beginning of the time step/period, transitions into the states 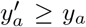 and 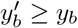, which can only become more infectious.
b. Develop Gaussian Mixture Bethe Free energy and Belief Propagation approaches for approximate inference of the S-Ising Models. The challenge here consists in constructing computationally efficient approximations for BP equations, which are functional integral equations. We are working on extending the approach of [62, 68] and approximate pair-wise and nodal probabilities, commonly called beliefs, via Gaussian Mixture. The Gaussian Mixture ansatz, once substituted in the Bethe Free Energy estimate for the log-partition function results in a finite dimensional but non-convex optimization. We intend to explore different ways to parameterize the Gaussian mixture to better represent integrals entering the Bethe Free Energy. We plan to design efficient algorithms to solve the resulting optimization problem providing estimates for the marginal probabilities of interest. We also intend to develop dual formulation consisting in the Gaussian Mixture substitute for messages (Lagrangian multipliers for constraints between pair-wise and nodal beliefs). Belief Propagation equations for messages will then be solved iteratively. We aim to juxtapose primal and dual approaches, possibly designing a novel primal-dual hybrid algorithm for resolving inference in the S-Ising Models.
c. Explore other (than based on the Gaussian Mixture) approaches to inference in the S-Ising Models. The alternatives may include piece-wise linear, or more generally polynomial, approximations of factor functions and finite dimensional regularizations of the S-Ising model. Piece-wise polynomial approximation of the factor functions naturally suggests to build variational algorithms with marginal beliefs, or messages (respective Lagrangian multipliers in the Bethe Free energy formulation) which are piece-wise polynomial as well. We will develop such algorithms, analyzing if the underlying functionals (for instance the Bethe Free energy) are convex and also testing if the newly developed algorithms provide upper-, lower-, or gap-guarantees. The M- (finite-) dimensional regularizations of the factor functions result in the models of the Potts type, where the Ising, (2 *×* 2), interaction matrix is replaced by the (*M × M*) matrix. Analyzing if the resulting *M*-state models remain attractive/supermodular (as the Ising model) will be our prime-focus. We will work on extending inference approaches, developed under auspecies of the Tasks #3c,d.
d. Consider generalizations of the S-Ising models accounting for multi-community (beyond pairwise) interactions, where factors are associated with larger clicks, such as triplet of communities, or pre-defined agglomeration of communities. Those multi-community interactions are coveted to model virus transmission at the public places, like airports, large malls, stores and related, attended by residents of multiple communities. As shown in [32], by constructing a by-partite GM connecting census tracts within the major metro-areas in USA to specific Points Of Interest (POI) – non residential locations that people visit such as restaurants, grocery stores and religious establishments - within the city and studying SEIR dynamics on the graph, a minority of POIs account for the majority of the predicted infections. We can build POI-GMs with each POIs explained in terms of a factor function dependent on the infection values of multiple census tracts contributing majority of visits to the POI. The factor GM will generalize the pair-wise GM discussed so far. We are interested to pose and solve inference problems over dynamic and static versions of the POI-GMs. Notice, that the so-called “Linear Threshold Model”, introduced and analyzed in the context of analysis and maximization/minimization of influence in social networks [74] (see also [118] and references therein), is an exemplary Dynamic GM of the factor graph type which is of interest to the epidemiology.

### E. Learning Graphical Models of Pandemic

Recall that we call the static Ising and S-Ising Models of Pandemic, as well as their dynamic counterparts, collectively the Graphical Models of Pandemic. So far we have assumed that the problem of statistical inference over the Graphical Models of Pandemic are well posed. For example, in the static case of the Ising and S-Ising models the assumption translates into the knowledge of (a) the underlying graph, (b) the pairwise interaction between communities/nodes, and (c) exogeneous and inhomogeneous infection biases of the communities. However, realistically, the Graphical Models may be known only through observed, or partially observed samples, and also through some additional specific information provided by the public health and transportation experts. This setting calls for the following “Learning Graphical Models” tasks:

**Task #5: Learning Graphical Models of Pandemic**. Aiming to approach modeling and mitigation reality of the actual pandemic, and especially of the COVID-19, in the most realistic way we intend to adapt the following Graphical Model Learning methodology to devising the Graphical Models of Pandemic.

a. The Graphical Models should be built based on the input of the public safety experts guiding the selection at the proper level of coarse-graining and quantifying the significance of nodal/community biases and pairwise interactions. This “expert”-informed input will be stated in terms of an a-priori choice of the specific sub-class of GM within the most general class of the Graphical Models considered, for example, it may result in identification of a sub-class of the S-Ising models. The sub-class may be formulated as fixing a graph, but leaving single-community biases and pair-wise community interactions uncertain. The selection of the sub-class may also result in keeping the graph itself largely uncertain except for pairwise interactions of central communities and remote nodes, such as links between downtown of Seattle and suburbs. We may also consider extending learning to a broader class of GM, e.g. bi-partite factor graph models representing higher-degree interactions at the public venues, such as places of worship, shopping malls, school, universities, etc. (See e.g. related discussion in Section VI D.)
b. When the class of possible Graphical Models is fixed we plan to apply the powerful GM learning approaches developed by the ML community. One may start with the classic in ML notion of the sufficient statistics [116]. This method requires (a) data-driven estimation of the sufficient statistics (in the case of the Ising model these are all first-order, i.e. nodal, moments, *∀a* ∈ 𝒱 : *m*_*a*_, and all second-order, or rather edge, moments, *∀*{*a, b*}∈ ℰ : 𝔼[*x*_*a*_*x*_*b*_]); (b) solving convex optimization of the Legendre-Fenchel type over the parameter tuple, (𝒢, ***J***, ***h***), which requires estimating the partition function, *Z*(𝒢, ***J***, ***h***), for each value of the tuple. Both of these steps are challenging, however for different reasons. Resolving (a) requires extracting information from the available epidemiological data. Recent impressive successes of many inter-disciplinary research teams in related tasks, see [32, 92], makes us hopeful that the hurdle can be overcome either based on the actual observational data or via synthetic data generated by a properly calibrated ABM. Bottleneck in implementing (b) is on the computational side and – we ought to rely on an efficient heuristics to resolve the inference problem (computing the partition function) discussed in Sections III B 2,VI C,VI D. An attractive “beyond sufficient statistics” approach consists in learning the GM of pandemics based on the pseudo-log-likelihood and/or interaction screening methodology developed recently. (See [87, 114] and references there in). We will also develop new hybrid methodologies adapting the aforementioned pseudo-log-likelihood and interaction screening approaches to specifics of the Graphical Models of Pandemic, for instance extending the techniques to learning DGM and broader class of static GM, for instance of the soft-Ising type discussed above in Section VI D.
c. Moreover, it will be of interest to pose and attempt to resolve an even more challenging question of “infernence” – combining inference and learning of the underlying Graphical Model in one optimization-based setting [66, 72]. We preset important inference questions, e.g. computing the probability for a surge triggered by a super-spreader event to result in infecting more than 10% of the **S**usceptible population in a block of communities, say around the Pike Market in Seattle. Then we intend to design Graphical Modeling algorithms aimed at the best quality of predictions for this specific inference problem (as opposed to many other inference question one may ask).

### F. Mitigating Pandemic

The many possible options for mitigating devastating effects of pandemics are split between pharmaceutical and non-pharmaceutical. The ultimate pharmaceutical option is vaccination, which recently became available in the World’s fight with the COVID-19 pandemic. However, and in spite of vaccine availability, there are number of challenges we need to resolve to put the pandemic under control, in particular

- Availability of vaccine is limited at least in the early period, especially in developing countries.
- Not all people will be taking vaccine.
- Efficiency of the vaccine depends on the version and it may be changing in time (for instance as the virus mutate) and it is by and large not known for certain population groups.
- It is not yet known whether vaccine immunity also guarantees halt of the immune individual’s transmissibility, or rather ability to infect others.
- Factors related to implementation will contribute more to the success of vaccination programs than a vaccine’s efficacy as determined in clinical trials [104].

All of the above suggest that the non-pharmaceutical options, currently in place around the globe to fight COVID-19, remain important. The non-pharmaceutical options include

- Enforcing social-distance and mask-wearing.
- Encouraging work from home, also creating other incentives to limit travel.
- Setting capacity limits or imposing closures, global or partial, of common places of social gathering such as schools, universities, restaurants, businesses, national parks, places of worship, and so on.
- Limiting air- and road-traffic, setting curfew of areas and roads, global or partial.

We expect that it will be possible to map some of the pharmaceutical and non-pharmaceutical actions listed above (and other actions which may become available as the fight with pandemics evolves, and our experience and resources grow) into nodes/communities local corrections of the respective components of the vector of nodal biases, ***h***, and other into corrections of the components of the inter community interaction vector, ***J***. For example, targeted vaccination of people residing at a census tract, *a*, decreases *h*_*a*_ (remind that large negative *h*_*a*_ means bias towards the healthy, **S**usceptable, state), while control of the traffic between the pair of census tracts, *a*, and, *b*, for instance achieved via road closures or curfews, decreases the value of the pair-wise interaction, *J*_*ab*_ *≥*0, possibly all the way to zero if the inter-node traffic is halted completely (we may also interpret it as cutting respective edge in the graph).

Any of the actions have a budget and other public health considerations which limit their availability and should be accounted for [1]. We may state the considerations formally as the vector of constraints, ***C***(𝒢, ***J***, ***h***) *≤* 0, imposed on the actionable parameters of the GM, or rather the vector of the pair-wise interactions, ***J***, and the vector of nodal biases, ***h***. Then, one plausible mitigation formulation may consists in minimizing the ALI-index (26) over the degrees of freedom which can be changed, subject to the constraints:

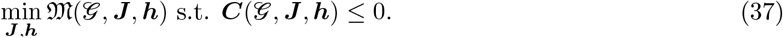

Other objectives, e.g. less global and targeting particular census tracts of a higher significance than others, may be stated similarly. Notice that translating actual actions, for instance traffic limitations, stay home orders, business closures, etc, into quantitative corrections to the current parameters of the GM, ***J*** and ***h***, will be needed for both expressing the public health guidance into the formal constraints in Eq. (37), and then also for mapping the optimal solution of Eq. (37) into actions (or rather actionable decision suggestions).

**Task #6: Mitigation of Pandemic Spread**.

a. **State the Pandemic Mitigation Problems** formally, e.g. in the minimal form of Eq. (37). The models should be based on the data availability, learning and prediction considerations discussed above. The main “new” challenge in building relevant mitigation formulations, i.e. the challenge specific to the mitigation/control formulation, will consists in translating informal language of regulations, recommendation and public health guidance into constraints and formulas. The models should be corroborated by the public health experts. Find existing or develop new solutions to the mitigation problems. The approaches may very dependent on details of the models, e.g. on if the control degrees of freedom are discrete or continuous and on complexity of the inner (inference) problem. One promising direction for research consists in reformulating the complex mitigation problem containing two (or more) levels as a combined optimization over all the (inference and control) degrees of freedom. In the following we discuss other relevant ideas focusing primarily on the cases where the mitigation/control degrees of freedom are geographical.
b. **Geographic Approaches: Formulations and Prior Studies**. During 2020, one of the tools that proved to be effective–though not necessarily popular in containing outbreaks was quarantining certain geographical regions so that people were either not allowed to cross in or out of these regions, or only allowed to do so within certain parameters (such as having a temperature less than or equal to 100^*°*^ F). Regions with a certain number of COVID-19 cases isolated themselves from regions with a significantly different, higher or lower number of cases. (The former case is more common in the US, while the latter is more common in Europe.) We will refer to this containment tool as regional quarantines. Once a quarantine is considered, many questions arise about the type of separation and its exact boundaries. The predicted impact of the quarantine, the resources in manpower required to enforce it, and the impact on the daily life of people living inside and outside the quarantined region are all closely tied to each other. Many undesired effects, including the risk of increasing the virus spread, might result from less-than-optimal decisions on the number of quarantined regions, their boundaries, and the type of movement the quarantine intends to limit. We aim to develop coherent systems that will process the geographic maps – known and predicted geographic constraints, patient information, mobility patterns and resource allocation such as food stores. Our system will draw proposed boundaries of optimal quarantined regions on a map. The term “optimal” here might refer to different user-specified objectives. Alternatively our system can evaluate proposed regional quarantine boundaries, and suggest their improvements. Considerations to be included in accessing the quarantine are: **Effectiveness** in minimizing the expected number of infected people. Disconnecting people from their resources, such as stores, forces people to look elsewhere, which might cause an undesirable stress on public infrastructure, or overcrowding of certain areas, therefore preventing proper social distancing. (In certain cases in Israel, emergency services had to carry food packages to multiple houses.) See Figure 9 for an illustration. **Accessing necessities** while planning the quarantine boundaries will minimize these effects. Quarantine regions should be easy to communicate and comprehend, and be associated with geographic boundaries, or rather be **compact**. Splitting Geographic regions should be avoided if possible. We will measure the region compactness as a combination of its boundary complexity and the *β*-stretchness [27] or-fatness [46] of the shape. The complexity measures the number of map segments on the boundary (where each map segment refers to a portion of a major street, or other geographic feature). Compactness might indicate that the resources needed to monitor the quarantine are minimized, as the ratio of the area to the squared boundary length is large, which in a sense, enforces short boundaries. Trade-offs in drawing quarantine boundaries are illustrated in the left panel of Fig. 9. We seek to back selection of boundaries with **mathematical guarantees**. Accepting quarantine, and determining its boundaries are, to the best of our knowledge, extremely controversial. We believe that discovering combinatorial properties, and mathematical proofs of the superiority of one solution with respect to the other, could help in this emotional debate. Notice, that the compactness argument above suggests that the boundaries will use major street, and that their number is small. Dimensionality of the example on the right panel of Fig. 9 is relatively small (8 vertices on the boundary of a cut). Hence even an exhaustive search over all such regions may not be prohibitively expensive. More complex boundaries can be drawn heuristically, e.g. utilizing Adaptive Belief Propagation [105] that might be adjusted to handle efficiently divide-and-conquer operations. (See discussion of other heuristics below.) The vast literature and prior studies in the fields of GIS and Computational Geometry, e.g. on facility location [25], the so-called stable marriage algorithm [47, 54] (that optimally assigns people to food stores while satisfying load factor and distances to the assigned store) should be utilized. The load factor is especially important in connections with the maintenance of the social distancing. We should aim at finding efficiently good quarantine boundaries while maintaining feasible matchings.
c. **Optimal Quarantines**. Let us list a number of viable principal solutions. **Clustering Boundaries and Compact Representation**. As shown in [28], we could maintain (implicitly) an exponential number of candidate paths, and assemble cycles from them, using only a relatively small number of clusters. We believe that these approaches will assist in maintaining the exponential number of possible paths. Such structures might be unavoidable, since in general the shortest weight-constrained path problem is NP-complete [55, page 214], and therefore approximations must be developed. **GM renormalization and VC-dimension**. The exponential cost of exhaustive search is computationally unbearable in a full-scale graph, but it can be tractable in much smaller graphs. Furthermore, we may also utilize the ideas of smart GM inference, discussed in the preceding Sections, and specifically exploring strategies of the GM reductions, such as mini-bucket renormalization [20, 21], to scan through the exponentially large family of the candidate paths efficiently. We plan to investigate how this and other GM reduction techniques can be used to find optimal quarantine cuts. Moreover, planarity or approximate planarity of the underlying GM may help. Indeed, as recently proven, certain combinatorial properties of planar graphs with respect to cuts (more technically, the ratio of pairs of vertices that are disconnected by a cut) can be *ε*-approximated by considering only vertex samples of surprisingly small size due to the finite Vapnik-Chervonenkis dimension [60] of the obtained sets of connected components. **Quarantine Cuts**. We will use the term “cut” to denote a single street, junction, or other geographic feature, where we intend to limit the traffic for the sake of quarantine. From the GM perspective, we associate a single cut with the binary operation of marking edges crossed by the cut. As resources are needed to quarantine the cut, a natural question becomes: what is the maximum benefit that we could reach by safeguarding at most *k* edges? It is known that a very restricted version of our problem is already NP-complete [55, page 210], which suggests to attack the problem with approximate methods and/or heuristics.
d. **Cut Heuristics**. Even much simpler geometric separation problems than these discussed so far are computationally hard [45, 50, 96]. A very popular and highly practical algorithmic technique to resolve computationally hard problems is an application of a sequence of greedy improvements to an initially chosen solution. (Municipal boundaries is a natural initial choice. Categorization of cities utilizing the “signal-light” monitoring system [14], adopted in the EU and other counties, may be another example of a reasonable initial choice. However, this initial guess might be sub-optimal in terms of reducing the virus spread, and might violate the considerations discussed above.) In the local search approach, we will improve it via local changes of the boundary, repeatedly replacing one boundary segments for another. Local improvements approach turned out to be instrumental in generating good partitions of geographical maps in the study of, the so-called partisan gerrymandering [23, 93], a problem that shares several components with our problem. We will improve the naÏve running time for finding an optimal cut with respect to the predicted number of infections based on the inference model of Section III for “almost” planar graphs, see Task #4b. Here, we consider a polynomial-size family of geometric separations, or less formally, simple geometries. For example, consider all cuts formed by a vertical line. There are *O*(*n*) combinatorially distinct locations of vertical lines in a graph with *n* nodes. Can we find the best line faster than in time *O*(*n*^5*/*3^)? The heart of the matter in this problem is the computation of the partition function. When computing the partition function of the underline GM, a cut in a graph introduces many pairs of independent variables. In this context, we intend to leverage the techniques of variable elimination such as bucket-elimination described in Section III. We also aim to explore **close-to-optimum quarantines** in the dynamic models, discussed in Section II and generalize it to a broader class of dynamic GM of pandemics. The success of this approach depends on whether sets of our elementary geographic cuts or their soft modifications exhibit a sub/supermodular or convex behavior [76, 77, 98]. Moreover, when optimizing the parameters of the quarantine boundaries, certain strong features such as the convexity or compactness of the boundaries of the quarantine regions, are also highly desirable as resulting in highly scalable algorithms. We aim to continue our pilot investigation on when and if the desirable property holds exactly or approximately in the dynamic GMs of pandemic.

## Data Availability

The data used in this manuscript are from the FLUTE, a publicly available stochastic influenza epidemic simulation model:
Dennis L Chao, M Elizabeth Halloran, Valerie J Obenchain, and Ira M Longini Jr. Flute, a publicly available stochastic influenza epidemic simulation model. PLoS Comput Biol, 6(1):e1000656, 2010

## Notes

### Competing Interest Statement

The authors have declared no competing interest.

### Funding Statement

This work is partially supported by NSF RAPID: Infer and Control Global Spread of Corona-Virus with Graphical Models

## References

[1] CDC center for Disease Control and Prevention, Implementation of Mitigation Strategies for Communities with Local COVID-19 Transmission. https://www.cdc.gov/coronavirus/2019-ncov/community/community-mitigation.html.

[2] CDC center for Disease Control and Prevention, Morbility and Mortality Weekly Reports. https://www.cdc.gov/mmwr/volumes/69/wr/mm6915e2.htm?s_cid=mm6915e2_x.

[3] Confirmed cases and deaths by country, territory, or conveyance, https://www.worldometers.info/coronavirus/\#countries.

[4] Covid-19 confirmed and forecasted case data. los alamos national laboratory. https://covid-19.bsvgateway.org/. xAccessed: 2010-10-26.

[5] Covid-19 global cases at johns hopkins university. https://gisanddata.maps.arcgis.com/apps/opsdashboard/index.html.

[6] Covid-19 scenarios. https://neherlab.org/covid19/.

[7] Gleamviz: Real time forecast of global epidemic. http://www.gleamviz.org/.

[8] Institute for health metrics & evaluation. http://www.healthdata.org/.

[9] MIDAS: Accessed: Models of Infectious Disease Agent Study, https://midasnetwork.us/.

[10] SafeGraph COVID-19 Data Consortium. Accessed: San Francisco, CA: SafeGraph Inc. https://www.safegraph.com/covid-19-data-consortium.

[11] Safegraph social distancing metrics. Accessed: san francisco, ca: Safegraph inc. https://docs.safegraph.com/docs/social-distancing-metrics.

[12] What about bias in safegraph data set? san francisco, Accessed: ca: Safegraph inc. https://www.safegraph.com/blog/what-about-bias-in-the-safegraph-dataset.

[13] Wiki template:2019–20 coronavirus pandemic data. https://en.wikipedia.org/wiki/Template:2019\%E2\%80\%9320_coronavirus_pandemic_data

[14] 2020.

[15] Adeel Afzal. Molecular diagnostic technologies for covid-19: Limitations and challenges. Journal of Advanced Research, 26:149–159, 2020.

[16] S. Ahn, M. Chertkov, and J. Shin. Synthesis of mcmc and belief propagation. In D. D. Lee, M. Sugiyama, U. V. Luxburg, I. Guyon, and R. Garnett, editors, Advances in Neural Information Processing Systems 29, pages 1453–1461. Curran Associates, Inc., 2016.

[17] S. Ahn, M. Chertkov, and J. Shin. Synthesis of MCMC and Belief Propagation. In D. D. Lee, M. Sugiyama, U. V. Luxburg, I. Guyon, and R. Garnett, editors, Advances in Neural Information Processing Systems 29, pages 1453–1461. Curran Associates, Inc., 2016.

[18] S. Ahn, M. Chertkov, and J. Shin. Gauging Variational Inference. In I. Guyon, U. V. Luxburg, S. Bengio, H. Wallach, R. Fergus, S. Vishwanathan, and R. Garnett, editors, Advances in Neural Information Processing Systems 30, pages 2885–2894. Curran Associates, Inc., 2017.

[19] S. Ahn, M. Chertkov, and J. Shin. Gauging Variational Inference. In I. Guyon, U. V. Luxburg, S. Bengio, H. Wallach, R. Fergus, S. Vishwanathan, and R. Garnett, editors, Advances in Neural Information Processing Systems 30, pages 2885–2894. Curran Associates, Inc., 2017.

[20] S. Ahn, M. Chertkov, J. Shin, and A. Weller. Gauged mini-bucket elimination for approximate inference. In A. J. Storkey and F. Pérez-Cruz, editors, International Conference on Artificial Intelligence and Statistics, AISTATS 2018, 9-11 April 2018, Playa Blanca, Lanzarote, Canary Islands, Spain, volume 84 of Proceedings of Machine Learning Research, pages 10–19. PMLR, 2018.

[21] S. Ahn, M. Chertkov, A. Weller, and J. Shin. Bucket renormalization for approximate inference. In J. G. Dy and A. Krause, editors, Proceedings of the 35th International Conference on Machine Learning, ICML 2018, Stockholmsmässan, Stockholm, Sweden, July 10-15, 2018, volume 80 of Proceedings of Machine Learning Research, pages 109–118. PMLR, 2018.

[22] Ethem Alpaydin. Introduction to machine learning. MIT press, 2020.

[23] Micah Altman, Michael P McDonald, et al. Bard: Better automated redistricting. Journal of Statistical Software. Forthcoming, URL http://www.jstatsoft.org, 42(4):p1–28, 2011.

[24] R.M. Anderson and R.M. May. Infectious Disease of Humans: Dynamics and Control. Oxford University Press, Oxford, 1991.

[25] Alireza Boloori Arabani and Reza Zanjirani Farahani. Facility location dynamics: An overview of classifications and applications. Computers & Industrial Engineering, 62(1):408–420, 2012.

[26] Julien Arino and Stéphanie Portet. A simple model for covid-19. Infectious Disease Modelling, 5:309–315, 2020.

[27] Esther Arkin, Alon Efrat, Cesim Erten, Ferran Hurtado, Joseph Mitchell, Valentin Polishchuk, and Carola Wenk. Shortest tour of a sequence of disjoint segments in l1. Proc. 16th Fall Workshop on Computational and Combinatorial Geometry, 2006.

[28] Esther M Arkin, Faryad Darabi Sahneh, Alon Efrat, Fabian Frank, Radoslav Fulek, Stephen Kobourov, and Joseph SB Mitchell. Computing β-stretch paths in drawings of graphs. In 17th Scandinavian Symposium and Workshops on Algorithm Theory (SWAT 2020). Schloss Dagstuhl-Leibniz-Zentrum für Informatik, 2020.

[29] F. Barahona. On the computational complexity of Ising spin glass models. Journal of Physics A: Mathematical and General, 15(10):3241, 1982.

[30] Martin Z. Bazant and John W. M. Bush. Beyond six feet: A guideline to limit indoor airborne transmission of covid-19. medRxiv, https://web.mit.edu/bazant/www/COVID-19/, 2020.

[31] Anass Bouchnita and Aissam Jebrane. A hybrid multi-scale model of covid-19 transmission dynamics to assess the potential of non-pharmaceutical interventions. Chaos, Solitons & Fractals, page 109941, 2020.

[32] Serina Chang, Emma Pierson, Pang Wei Koh, Jaline Gerardin, Beth Redbird, David Grusky, and Jure Leskovec. Mobility network models of COVID-19 explain inequities and inform reopening. Nature, November 2020.

[33] Dennis L Chao, M Elizabeth Halloran, Valerie J Obenchain, and Ira M Longini Jr. Flute, a publicly available stochastic influenza epidemic simulation model. PLoS Comput Biol, 6(1):e1000656, 2010.

[34] Shizhe Chen, Daniela M Witten, and Ali Shojaie. Selection and estimation for mixed graphical models. Biometrika, 102(1):47–64, March 2015. Edition: 2014/12/24.

[35] M. Chertkov, R. Abrams, A. Esmaieeli, and A. Efrat. Graphical models of pandemic. In preparation, 2020.

[36] M. Chertkov and V.Y. Chernyak. Loop calculus in statistical physics and information science. Phys. Rev. E, 73:065102, 2006.

[37] Michael Chertkov, Vladimir Chernyak, and Yury Maximov. Gauges, Loops, and Polynomials for Partition Functions of Graphical Models. arXiv:1811.04713, 2018.

[38] Michael Chertkov and Vladimir Y Chernyak. Loop series for discrete statistical models on graphs. Journal of Statistical Mechanics: Theory and Experiment, 2006(06):P06009–P06009, 2006.

[39] Phillip C Cooley, D Roberts, VD Bakalov, SBikmal, S Cantor, T Costandine, L Ganapathi, BJ Golla, G Grubbs, C Hollingsworth, S Li, Y Qin William Savage, D Simoni E Solano, and D Wagener. The model repository of the models of infectious disease agent study. IEEE transactions on information technology in biomedicine : a publication of the IEEE Engineering in Medicine and Biology Society, 12(4):513–522, July 2008. Publisher: IEEE.

[40] Rina Dechter. Bucket elimination: A unifying framework for reasoning. Artificial Intelligence, 113(1):41–85, 1999.

[41] Rina Dechter and Irina Rish. Mini-buckets: A general scheme for bounded inference. J. ACM, 50(2):107–153, March 2003.

[42] Paul L Delamater, Erica J Street, Timothy F Leslie, Y Tony Yang, and Kathryn H Jacobsen. Complexity of the Basic Reproduction Number (R(0)). Emerging infectious diseases, 25(1):1–4, January 2019. Publisher: Centers for Disease Control and Prevention.

[43] Josip Djolonga and Andreas Krause. From map to marginals: Variational inference in bayesian submodular models. In Advances in Neural Information Processing Systems, pages 244–252, 2014.

[44] Allen Downey. Think Complexity: Complexity Science and Computational Modeling. O’Reilly Media, Inc., 2nd edition, 2018.

[45] Peter Eades and David Rappaport. The complexity of computing minimum separating polygons. Pattern Recognition Letters, 14(9):715–718, 1993.

[46] Alon Efrat and Micha Sharir. On the complexity of the union of fat convex objects in the plane. Discrete & Computational Geometry, 23(2):171–189, 2000.

[47] David Eppstein, Michael T Goodrich, Doruk Korkmaz, and Nil Mamano. Defining equitable geographic districts in road networks via stable matching. In Proceedings of the 25th ACM SIGSPATIAL International Conference on Advances in Geographic Information Systems, pages 1–4, 2017.

[48] S. Eubank, I. Eckstrand, B. Lewis, S. Venkatramanan, M. Marathe, and C. L. Barrett. Commentary on ferguson, et al., “impact of non-pharmaceutical interventions (NPIs) to reduce COVID-19 mortality and healthcare demand”. 82(4):52.

[49] Stephen Eubank, Hasan Guclu, V. S. Anil Kumar, Madhav V. Marathe, Aravind Srinivasan, Zoltán Toroczkai, and Nan Wang. Modelling disease outbreaks in realistic urban social networks. Nature, 429(6988):180–184, May 2004.

[50] Sandor Fekete. On the complexity of min-link red-blue separation. Manuscript, 1992.

[51] Neil Ferguson, Daniel Laydon, Gemma Nedjati-Gilani, Natsuko Imai, Kylie Ainslie, Marc Baguelin, Sangeeta Bhatia, Adhiratha Boonyasiri, Zulma M. Cucunubá, Gina Cuomo-Dannenburg, Amy Dighe, Ilaria Dorigatti, Han Fu, Katy Gaythorpe, Will Green, Arran Hamlet, Wes Hinsley, Lucy Okell, Sabine van Elsland, and Azra Ghani. Report 9: Impact of non-pharmaceutical interventions (npis) to reduce covid-19 mortality and healthcare demand. 2020.

[52] Neil M. Ferguson, Derek A. T. Cummings, Christophe Fraser, James C. Cajka, Philip C. Cooley, and Donald S. Burke. Strategies for mitigating an influenza pandemic. Nature, 442(7101):448–452, July 2006.

[53] Neil M. Ferguson, Derek A.T. Cummings, Simon Cauchemez, Christophe Fraser, Steven Riley, Aronrag Meeyai, Sopon Iamsirithaworn, and Donald S. Burke. Strategies for containing an emerging influenza pandemic in Southeast Asia. Nature, 437(7056):209–214, September 2005.

[54] David Gale and Lloyd S Shapley. College admissions and the stability of marriage. The American Mathematical Monthly, 69(1):9–15, 1962.

[55] M. R. Garey and D. S. Johnson. Computers and Intractability: A Guide to the Theory of NP-Completeness. W. H. Freeman, 1979.

[56] Timothy C. Germann, Kai Kadau, Ira M. Longini, and Catherine A. Macken. Mitigation strategies for pandemic influenza in the united states. Proceedings of the National Academy of Sciences, 103(15):5935–5940, 2006.

[57] A. Globerson and T.S. Jaakkola. Approximate inference using planar graph decomposition. In Advances in Neural Information Processing Systems, pages 473–480, 2007.

[58] V. Gómez, H.J. Kappen, and M. Chertkov. Approximate inference on planar graphs using loop calculus and belief propagation. J. Mach. Learn. Res., 11:1273–1296, 2010.

[59] Manuel Gomez-Rodriguez, Jure Leskovec, and Andreas Krause. Inferring networks of diffusion and influence. ACM Trans. Knowl. Discov. Data, 5(4), February 2012.

[60] Guy Grebla, Alon Efrat, Esther Ezra, Rom Pinchasi, and Swaminathan Sankararaman. Data recovery after geographic correlated attacks. In 11th International Conference on the Design of Reliable Communication Networks, DRCN 2015, Kansas City, MO, USA, March 24-27, 2015, pages 65–72, 2015.

[61] C.G. Grijalva, M.A. Rolfes, Y. Zhu, and et al. Transmission of sars-cov-2 infections in households — tennessee and wisconsin, http://dx.doi.org/10.15585/mmwr.mm6944e1. MMWR Morb Mortal Wkly Rep. ePub, 2020.

[62] Yuanzhen Guo, Hao Xiong, and Nicholas Ruozzi. Marginal inference in continuous markov random fields using mixtures. Proceedings of the AAAI Conference on Artificial Intelligence, 33:7834–7841, 07 2019.

[63] M. Elizabeth Halloran, Neil M. Ferguson, Stephen Eubank, Ira M. Longini, Derek A. T. Cummings, Bryan Lewis, Shufu Xu, Christophe Fraser, Anil Vullikanti, Timothy C. Germann, Diane Wagener, Richard Beckman, Kai Kadau, Chris Barrett, Catherine A. Macken, Donald S. Burke, and Philip Cooley. Modeling targeted layered containment of an influenza pandemic in the united states. Proceedings of the National Academy of Sciences, 105(12):4639–4644, 2008.

[64] A. Hartmann and H. Rieger. New Optimization Algorithms in Physics. Wiley-VCH, 2004.

[65] Jonas M. B. Haslbeck and Lourens J. Waldorp. Estimating time-varying mixed graphical models in highdimensional data. Journal of Statistical Software, 93(8):1–46, 2020.

[66] U. Heinemann and A. Globerson. Inferning with high girth graphical models. ICML, 2014.

[67] Herbert W. Hethcote. The mathematics of infectious diseases. SIAM Rev., 42(4):599–653, December 2000.

[68] Marco F. Huber, Tim Bailey, Hugh Durrant-Whyte, and Uwe D. Hanebeck. On entropy approximation for gaussian mixture random vectors. 10 2008.

[69] Ayaz Hyder, David L. Buckeridge, and Brian Leung. Predictive validation of an influenza spread model. PLOS ONE, 8(6):1–20, 06 2013.

[70] M. Jerrum and A. Sinclair. Polynomial-time approximation algorithms for the ising model. SIAM Journal on Computing, 22(5):1087–1116, 1993.

[71] Jason K. Johnson, Vladimir Y. Chernyak, and Michael Chertkov. Orbit-product representation and correction of gaussian belief propagation. In Proceedings of the 26th Annual International Conference on Machine Learning, ICML ‘09, pages 473–480, New York, NY, USA, 2009. ACM.

[72] M. Karzand and G. Bresler. Inferning trees. In 2015 53rd Annual Allerton Conference on Communication, Control, and Computing (Allerton), pages 1344–1351, 2015.

[73] Efthimios Kaxiras and Georgios Neofotistos. Multiple epidemic wave model of the covid-19 pandemic: Modeling study. Journal of Medical Internet Research, 22(7):e20912, 2020.

[74] David Kempe, Jon Kleinberg, and Éva Tardos. Maximizing the spread of influence through a social network. In Proceedings of the Ninth ACM SIGKDD International Conference on Knowledge Discovery and Data Mining, KDD ’03, page 137–146, New York, NY, USA, 2003. Association for Computing Machinery.

[75] William Ogilvy Kermack, A. G. McKendrick, and Gilbert Thomas Walker. A contribution to the mathematical theory of epidemics. Proceedings of the Royal Society of London. Series A, Containing Papers of a Mathematical and Physical Character, 115(772):700–721, 1927.

[76] Elias Khalil, Bistra Dilkina, and Le Song. Cuttingedge: influence minimization in networks. In Proceedings of Workshop on Frontiers of Network Analysis: Methods, Models, and Applications at NIPS, 2013.

[77] Elias B. Khalil, Bistra Dilkina, and Le Song. Cuttingedge: Influence minimization in networks. In Workshop on Frontiers of Network Analysis: Methods, Models, and Applications at NIPS, 2013.

[78] D. Koller and N. Friedman. Probabilistic Graphical Models. Massachusetts: MIT Press, 2009.

[79] Vladimir Kolmogorov and Martin J. Wainwright. On the optimality of tree-reweighted max-product message-passing. In Proceedings of the Twenty-First Conference on Uncertainty in Artificial Intelligence, UAI’05, page 316–323, Arlington, Virginia, USA, 2005. AUAI Press.

[80] Samuel Lalmuanawma, Jamal Hussain, and Lalrinfela Chhakchhuak. Applications of machine learning and artificial intelligence for covid-19 (sars-cov-2) pandemic: A review. Chaos, Solitons & Fractals, 139:110059, 2020.

[81] Bruce Y Lee, Shawn T Brown, Philip C Cooley, Richard K Zimmerman, William D Wheaton, Shanta M Zimmer, John J Grefenstette, Tina-Marie Assi, Timothy J Furphy, Diane K Wagener, et al. A computer simulation of employee vaccination to mitigate an influenza epidemic. American journal of preventive medicine, 38(3):247–257, 2010.

[82] Jason D Lee and Trevor J Hastie. Learning the Structure of Mixed Graphical Models. Journal of computational and graphical statistics : a joint publication of American Statistical Association, Institute of Mathematical Statistics, Interface Foundation of North America, 24(1):230–253, January 2015.

[83] V. Likhosherstov, Y. Maximov, and M. Chertkov. Inference and sampling of k33-free Ising models. arXiv:1812.09587, 2018.

[84] V. Likhosherstov, Y. Maximov, and M. Chertkov. A new family of tractable Ising models. arXiv:1906.06431, 2019.

[85] V. Likhosherstov, Y. Maximov, and M. Chertkov. Tractable minor-free generalization of planar zero-field Ising models. arXiv:1910.11142, 2019.

[86] Qiang Liu and A. Ihler. Bounding the partition function using holder’s inequality. In ICML, 2011.

[87] A.Y. Lokhov, M. Vuffray, S. Misra, and M. Chertkov. Optimal structure and parameter learning of ising models. Science Advances, 4(3), 2018.

[88] Ira M. Longini, Azhar Nizam, Shufu Xu, Kumnuan Ungchusak, Wanna Hanshaoworakul, Derek A T Cummings, and M. Elizabeth Halloran. Containing pandemic influenza at the source. Science, 309(5737):1083–1087, August 2005.

[89] Gina Lovasi and et.al. Population health methods: Agent based modeling, 2020.

[90] Maimuna Majumder and Kenneth D. Mandl. Early transmissibility assessment of a novel coronavirus in wuhan, china (january 26, 2020). In SSRN.

[91] Dmitry M. Malioutov, Jason K. Johnson, and Alan S. Willsky. Walk-sums and belief propagation in gaussian graphical models. J. Mach. Learn. Res., 7:2031–2064, December 2006.

[92] Sergei Maslov and Nigel Goldenfeld. Flattening the curve fails unless done very early: results from a simulation of icu capacity in chicago, 2020.

[93] Jonathan C Mattingly and Christy Vaughn. Redistricting and the will of the people. arXiv preprint arXiv:1410.8796, 2014.

[94] Mariusz Maziarz and Martin Zach. Agent-based modelling for sars-cov-2 epidemic prediction and intervention assessment: A methodological appraisal. Journal of Evaluation in Clinical Practice, 26(5):1352–1360, 2020.

[95] C. Merow and M. C. Urban. Seasonality and uncertainty in global covid-19 growth rates. Proceedings of the National Academy of Sciences, 2020.

[96] Joseph S.B. Mitchell and Subhash Suri. Separation and approximation of polyhedral objects. Computational Geometry, 5(2):95–114, 1995.

[97] K. Murphy. Machine Learning: A Probabilistic Perspective. Massachusetts: MIT Press, 2011.

[98] Mukund Narasimhan and Jeff Bilmes. A submodular-supermodular procedure with applications to discriminative structure learning. In Proceedings of the Twenty-First Conference on Uncertainty in Artificial Intelligence, UAI’05, page 404–412. AUAI Press, 2005.

[99] Radford M. Neal. Probabilistic inference using markov chain monte carlo methods. Technical report, 1993.

[100] Praneeth Netrapalli and Sujay Sanghavi. Learning the graph of epidemic cascades. In Proceedings of the 12th ACM SIGMETRICS/PERFORMANCE Joint International Conference on Measurement and Modeling of Computer Systems, SIGMETRICS ‘12, page 211–222, New York, NY, USA, 2012. Association for Computing Machinery.

[101] Even the coauthor CNP Slagle and his immediate family have suffered greatly from the illness.

[102] This data has recently been used by the Centers for Disease Control and Prevention to gather information on the degree to which social distancing has been predictive in the United States following the COVID-19 outbreak [2]. Notice also, that analyses by SafeGraph show that their sample correlates very highly with the true Census populations [0.97 across U.S. counties] and with the proportion of the population across educational attainment and income levels [correlation of 0.99]. See: [12].

[103] OpenStreetMap contributors. Planet dump retrieved from https://planet.osm.org, 2017.

[104] A. David Paltiel, Jason L. Schwartz, Amy Zheng, and Rochelle P. Walensky. Clinical outcomes of a covid-19 vaccine: Implementation over efficacy. Health Affairs, 0(0):10.1377/hlthaff.2020.02054, 0. PMID: 33211536.

[105] Georgios Papachristoudis and John W. Fisher. Adaptive belief propagation. In Proceedings of ICML, 2015.

[106] Tomas Pueyo. Coronavirus: The hammer and the dance. https://tomaspueyo.medium.com/coronavirus-the-hammer-and-the-dance-be9337092b56.

[107] Robert C. Reiner, Ryan M. Barber, James K. Collins, Peng Zheng, Christopher Adolph, James Albright, Catherine M. Antony, Aleksandr Y. Aravkin, Steven D. Bachmeier, Bree Bang-Jensen, Marlena S. Bannick, Sabina Bloom, Austin Carter, Emma Castro, Kate Causey, Suman Chakrabarti, Fiona J. Charlson, Rebecca M. Cogen, Emily Combs, Xiaochen Dai, William James Dangel, Lucas Earl, Samuel B. Ewald, Maha Ezalarab, Alize J. Ferrari, Abraham Flaxman, Joseph Jon Frostad, Nancy Fullman, Emmanuela Gakidou, John Gallagher, Scott D. Glenn, Erik A. Goosmann, Jiawei He, Nathaniel J. Henry, Erin N. Hulland, Benjamin Hurst, Casey Johanns, Parkes J. Kendrick, Apurva Khemani, Samantha Leigh Larson, Alice Lazzar-Atwood, Kate E. LeGrand, Haley Lescinsky, Akiaja Lindstrom, Emily Linebarger, Rafael Lozano, Rui Ma, Johan Månsson, Beatrice Magistro, Ana M. Mantilla Herrera, Laurie B. Marczak, Molly K. Miller-Petrie, Ali H. Mokdad, Julia Deryn Morgan, Paulami Naik, Christopher M. Odell, James K. O’Halloran, Aaron E. Osgood-Zimmerman, Samuel M. Ostroff, Maja Pasovic, Louise Penberthy, Geoffrey Phipps, David M. Pigott, Ian Pollock, Rebecca E. Ramshaw, Sofia Boston Redford, Grace Reinke, Sam Rolfe, Damian Francesco Santomauro, John R. Shackleton, David H. Shaw, Brittney S. Sheena, Aleksei Sholokhov, Reed J. D. Sorensen, Gianna Sparks, Emma Elizabeth Spurlock, Michelle L. Subart, Ruri Syailendrawati, Anna E. Torre, Christopher E. Troeger, Theo Vos, Alexandrea Watson, Stefanie Watson, Kirsten E. Wiens, Lauren Woyczynski, Liming Xu, Jize Zhang, Simon I. Hay, Stephen S. Lim, Christopher J. L. Murray, and IHME COVID-19 Forecasting Team. Modeling COVID-19 scenarios for the United States. Nature Medicine, October 2020.

[108] Nir Rosenfeld, Mor Nitzan, and Amir Globerson. Discriminative learning of infection models. In Proceedings of the Ninth ACM International Conference on Web Search and Data Mining, WSDM ’16, page 563–572, New York, NY, USA, 2016. Association for Computing Machinery.

[109] R. Ross. The Prevention of Malaria. John Murray, London, 1910.

[110] M. I. Schlesinger. Syntatic analysis of two-dimensional visual signals in noisy conditions. Kibernetika [in Russian], 4:113–130, 1976.

[111] Alessandro Sette and Shane Crotty. Pre-existing immunity to SARS-CoV-2: the knowns and unknowns. Nature Reviews Immunology, 20(8):457–458, August 2020.

[112] Flaminio Squazzoni, J. Gareth Polhill, Bruce Edmonds, Petra Ahrweiler, Patrycja Antosz, Geeske Scholz, Emile Chappin, Melania Borit, Harko Verhagen, Francesca Giardini, and Nigel Gilbert. Computational models that matter during a global pandemic outbreak: A call to action. Journal of Artificial Societies and Social Simulation, 23(2):10, 2020.

[113] Devi Sridhar and Maimuna S Majumder. Modelling the pandemic. BMJ, 369, 2020.

[114] M. Vuffray, S. Misra, A. Lokhov, and M. Chertkov. Interaction Screening: Efficient and Sample-Optimal Learning of Ising Models. In D. D. Lee, M. Sugiyama, U. V. Luxburg, I. Guyon, and R. Garnett, editors, Advances in Neural Information Processing Systems 29, pages 2595–2603. Curran Associates, Inc., 2016.

[115] Philip V’kovski, Annika Kratzel, Silvio Steiner, Hanspeter Stalder, and Volker Thiel. Coronavirus biology and replication: implications for SARS-CoV-2. Nature Reviews Microbiology, October 2020.

[116] M.J. Wainwright and M.I. Jordan. Graphical models, exponential families, and variational inference. Foundations and Trends in Machine Learning, 1(1):1–305, 2008.

[117] Liang Wang, Xavier Didelot, Jing Yang, Gary Wong, Yi Shi, Wenjun Liu, George F. Gao, and Yuhai Bi. Inference of person-to-person transmission of COVID-19 reveals hidden super-spreading events during the early outbreak phase. Nature Communications, 11(1):5006, October 2020.

[118] Xinjue Wang, Ke Deng, Jianxin Li, Jeffery Xu Yu, Christian S Jensen, and Xiaochun Yang. Efficient targeted influence minimization in big social networks. World Wide Web, pages 1–18, 2020.

[119] T. Werner. A linear programming approach to max-sum problem: A review. Pattern Analysis and Machine Intelligence, IEEE Transactions on, 29(7):1165–1179, July 2007.

[120] Wikipedia. Agent Based Models, 2020.

[121] J. S. Yedidia, W. T. Freeman, and Y. Weiss. Constructing free-energy approximations and generalized belief propagation algorithms. IEEE Transactions on Information Theory, 51(7):2282–2312, 2005.

